# Maternal pre-pregnancy body mass index and risk of preterm birth: a collaboration using large routine health datasets

**DOI:** 10.1101/2023.04.12.23288473

**Authors:** RP Cornish, MC Magnus, SK Urhoj, G Santorelli, LG Smithers, D Odd, A Fraser, SE Håberg, A-M Nybo Andersen, K Birnie, JW Lynch, K Tilling, DA Lawlor

**Affiliations:** Population Health Sciences at the University of Bristol Medical School, University of Bristol, Oakfield House, Oakfield Road, Bristol, BS8 2BN, UK; MRC Integrative Epidemiology Unit at the University of Bristol, UK; Centre for Fertility and Health, Norwegian Institute of Public Health, Norway; Department of Public Health, Faculty of Health Sciences, University of Copenhagen, Denmark; Bradford Institute for Health Research, Bradford Royal Infirmary, UK; School of Public Health, University of Adelaide, Australia; School of Health and Society, University of Wollongong, Australia; Division of Population Medicine, Cardiff University School of Medicine

**Author notes:** These authors contributed equally (K Tilling, DA Lawlor).

## Abstract

**Importance:** Preterm birth (PTB), is a leading cause of child morbidity and mortality. Objective: To examine the associations of maternal pre-pregnant body mass index (BMI) with any PTB, spontaneous (SPTB) and medically indicated PTB (MPTB).

**Design:** A meta-analysis of eight population-based datasets.

**Setting:** Three UK datasets, two USA datasets, and one each from South Australia, Norway and Denmark, with different characteristics and sources of bias.

**Participants:** All pregnancies resulting in a live birth or stillbirth after 24 completed gestational weeks.

**Exposure:** Maternal pre-or early pregnancy BMI derived from self-reported or measured weight and height between 12 months pre-pregnancy and 15 weeks gestation.

**Main Outcome(s) and Measures(s):** Any PTB (delivery <37 completed weeks), SPTB and medically indicated PTB. Fractional polynomial multivariable logistic regression was applied to eight datasets from different high-income countries and time periods. The results were combined using a random effects meta-analysis.

**Results:** We found non-linear associations between pre-pregnant BMI and all three outcomes, across all datasets. The adjusted risk of any PTB and MPTB was elevated at both low and high BMIs, whereas the risk of SPTB was increased at lower levels of BMI but remained low or increased only slightly with higher BMI. In the meta-analysed data, the lowest risk of any PTB was at a BMI of 24.5 kg/m^2^ (95% confidence interval: 23.1, 30.3), with a value of 21.3 kg/m^2^ (20.8, 21.9) for MPTB; for SPTB, the risk remained roughly constant above a BMI of around 25-30 kg/m^2^.

**Conclusions and Relevance:** Consistency of findings across different populations, despite differences between them in the time period covered, BMI distribution, missing data and control for key confounders, highlight the importance of promoting pre-conception BMI between 21 to 30 kg/m2 to prevent MPTB and SPTB

## Introduction

Preterm birth (PTB; birth before 37 completed weeks gestation) affects around 10% of pregnancies worldwide. It is the leading cause of perinatal mortality and morbidity, and of childhood death up to 5 years^1,2^. Recent increases in PTB^2,3^ may be related to the obesity epidemic.

PTB can be medically indicated (MPTB) or spontaneous (SPTB). MPTB is driven by obstetric interventions (induction of labour or planned caesarean section) related to pregnancy complications such as pre-eclampsia or gestational diabetes and thus may be higher in women who are overweight or obese^4^. While the detrimental effects of MPTB is a trade off with detrimental effects of continued pregnancy in the presence of such conditions, SPTB is a major concern obstetrically because of its unpredictable nature.

Evidence from systematic reviews suggests an increased risk of PTB with both maternal overweight/obesity and underweight^5-10^, with some studies suggesting underweight might be a greater factor than obesity in SPTB^5-7,11-14^. Previous studies have largely explored established BMI categories and not attempted to identify the BMI with lowest risk or compared associations across countries with different levels of obesity. Our aim was to compare associations of maternal BMI with PTB, SPTB and MPTB across populations with differing characteristics, and to identify the optimal BMI with lowest risk for these outcomes.

## Methods

### Datasets

The datasets are described in Figure 1, summarising differences in key characteristics such as: years covered; BMI distribution; availability of confounders; and data completeness, with further details regarding missing data given in Supplementary Figure S1. Datasets ranged in size from just under 5,000 to over 23 million pregnancies and included three UK, two US, and one dataset each from Australia, Norway and Denmark. The datasets from Norway and Denmark and one from the USA (US vital statistics data) included all registered births across the countries during the study period. The years covered varied, with all except one (Collaborative Perinatal Project, USA, 1959-1965) including recent data.

**Figure 1.**
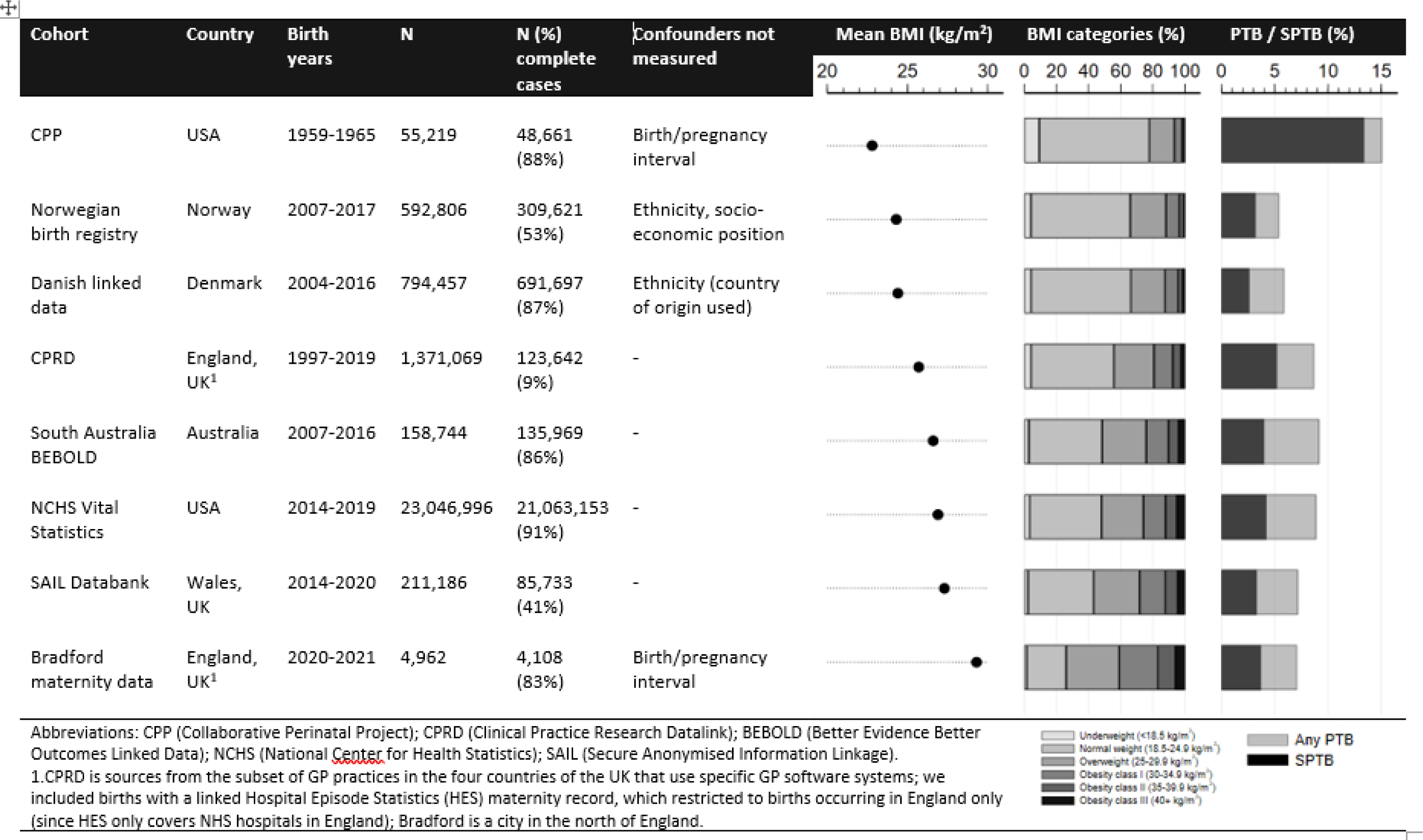
Key characteristics of the datasets

### Collaborative Perinatal Project (CPP)

CPP recruited just over 46,000 women with 59,391 pregnancies from twelve US centres providing prenatal care between 1959 and 1965. Socio-demographic, behavioural and physical data were collected at prenatal visits^15,16^.

### Norwegian birth registry

We used data between 2008 and 2017 from the Birth Registry of Norway (MBRN), which includes mandatory registrations of all pregnancies in Norway ending after 12 completed gestational weeks^17^.

### Danish linked data

We included all live births and stillbirths in Denmark between 2004 and 2016, using linked information from the Danish Medical Birth Registry^18^ and population registers held by Statistics Denmark.

### Clinical Practice Research Datalink (CPRD)

CPRD is a population-based database of primary care data from across the UK^19^ linked to other datasets. We included all pregnancies from the CPRD (GOLD) Pregnancy Register^20^ resulting in a live or still birth and with a linked record in the Hospital Episode Statistics (HES) maternity data; the latter covers NHS hospitals in England only.

South Australian Better Evidence Better Outcomes Linked Data (BEBOLD) platform Pregnancy data was obtained from the BEBOLD platform, which includes the South Australian Perinatal Statistics Collection 2007-2016, a mandatory collection of all births at least 400 grams or 20 weeks gestation^21^.

### US National Center for Health Statistics Vital Statistics (NCHS) data

We used publicly available birth and fetal death datasets from 2014 to 2019. These include information from mandatory registrations of all births and fetal deaths; for most states this includes fetal deaths of at least 350g and/or 20 weeks gestation^22^.

### Secure Anonymised Information Linkage (SAIL) Databank

The SAIL databank contains de-identified health and administrative data on the population of Wales, UK. We included pregnancies resulting in a live or still birth from 2014 onwards with a birth record in either the Maternity Indicators Dataset (MID)^23^ (data from the first antenatal assessment plus labour and birth) or the National Community Health (NCCH) database (birth registration and other data).

### Bradford maternity data

Maternity record data for all births at Bradford Royal Infirmary (BRI) between January 2020 and March 2021 were obtained from BRI Informatics Department.

Further details of each dataset are provided in supplementary materials.

### Outcomes

The primary outcome measures were any PTB, SPTB (delivery <37 completed weeks, with spontaneous onset of labour) and MPTB (labour induced or delivery initiated by caesarean section prior to onset of labour). Fetal deaths occurring up to 23 weeks, 6 days of gestation were excluded since these were absent or incomplete in most datasets. Secondary outcomes were very PTB, SPTB and MPTB (<32 weeks). Gestational age at delivery was predominantly based on early ultrasound measurements except in the CPP, where it was calculated from last menstrual period (LMP) (details in supplementary materials).

### Exposure

The exposure was maternal pre- or early pregnancy BMI, calculated from self-reported or measured pre-pregnancy or early pregnancy weight and height (details in supplementary materials).

### Covariates

The following confounders were identified a priori^24^: maternal age at birth, parity, ethnicity, smoking, socio-economic position (SEP), and birth or pregnancy interval. The availability of these confounders varied, as summarised in Figure 1 and with further details in supplementary materials. Because of its strong association with preterm birth, pregnancy size (singleton/multiple) was included as a covariate to increase precision. We decided a priori to maximise confounder adjustment within each dataset by not harmonising variables across datasets (where recorded differently or unavailable) but using the most detailed measures within each.

### Statistical methods

All analyses were carried out with pregnancy as the unit of analysis. For the primary analysis, multivariable logistic regression using fractional polynomials^25^ with up to three powers of BMI was used to examine the association between BMI and any PTB, SPTB, and MPTB. Datasets where mothers had more than one recorded pregnancy used robust standard errors if there was a unique mother ID variable. For each outcome, we chose an optimal model (in terms of the fractional polynomial; all models included all available confounders) that fit well in all datasets (and was potentially the best fitting model in several). Once the optimal model for each outcome had been selected, we carried out a multivariate, random effects meta-analysis with inverse variance weighting on the aggregate data. It was not possible to combine individual-level data, as most datasets had to be analysed on secure servers in different locations. Confounder-adjusted risks of any PTB, MPTB and SPTB were calculated from the optimal model and plotted against BMI. Where possible, the same reference group was used and consisted of singleton pregnancies, nulliparous, maternal age 25-29 years, non-smoker, pregnancy/birth interval not < 12 months, White/Caucasian. For SEP, which was measured in various ways, the reference category was the median group. In the Danish dataset, where country of origin was measured but not ethnicity, originating from Denmark was the reference group. Where feasible, the estimated BMI at which the risk of each outcome was lowest was calculated via differentiation and a 95% confidence interval (CI) obtained using bootstrapping (details in supplementary materials).

We conducted two secondary analyses. Firstly, we used standard WHO BMI categories (underweight <18.5 kg/m^2^, healthy weight 18.5-24.9, overweight 25-29.9, obesity class I 30-34.9, obesity class II 35-39.9, obesity class III 40+), to enable our results to be compared to other publications. Secondly, we examined very PTB (<32 completed weeks gestation), as this is related to more adverse outcomes than births from 33 to <37 weeks^26^.

Various sensitivity analyses were conducted. Firstly, to account for the fact that a woman with a MPTB could not have a SPTB and vice versa, models for SPTB were weighted by the inverse of one minus the probability of being a MPTB; conversely, models for MPTB were weighted by the inverse of one minus the probability of being a SPTB. The models for the weights included the same variables as the analysis model. Secondly, we carried out analyses excluding (i) stillbirths, (ii) post term deliveries (≥42 completed weeks gestation) and (iii) multiple births. Finally, because the CPP was carried out in the 1960s, with all other datasets contributing recent data, reflecting more contemporary practice and monitoring, we repeated the meta-analyses excluding this dataset.

We hypothesised that, within datasets, some covariates – particularly BMI and ethnicity – might be missing not at random (specifically, less likely to be missing if individuals had either a high or low BMI, or were not from an ethnic minority group). Thus, we decided a priori to use a complete case analysis in all datasets because in this situation (covariates missing not at random), multiple imputation would produce bias, whereas a complete case logistic regression gives unbiased estimates unless the chance of being a complete case depends on both the exposure and outcome^27,28^, which we thought unlikely.

All analyses were carried out in Stata; meta-analysis used Stata’s mvmeta command^29^.

## Results

Between 68% (CPRD) and 100% (South Australian BEBOLD) of pregnancies had gestational age at delivery recorded. In most datasets, BMI had the most missing data and between 9% (CPRD) and 92% (US Vital Statistics) were complete cases (Figure 1; Supplementary Figure S1). In CPRD, because BMI came from primary care data, thus relying on weight to have been measured near the time of conception as part of routine care, this information was only available for a small proportion of pregnancies. Supplementary Tables S1 to S8 give characteristics of the whole sample and complete cases for each dataset; across all datasets, characteristics were similar.

The risks of PTB and SPTB among complete cases were lowest in the Norwegian birth registry (5.4% PTB, 2.6% SPTB) and highest in the Collaborative Perinatal Project (CPP) (15.2% and 13.4%, respectively) (Figure 1). The risk of MPTB ranged from 1.6% (CPP) to 4.9% (US Vital Statistics). In CPP, SPTB accounted for around 90% of PTB; in the other datasets it ranged from 43% to 58%. The mean BMI ranged from 22.8 kg/m^2^ (CPP) to 29.3 kg/m^2^ (Bradford) (Figure 1; Supplementary Tables S1-S8).

### Main results

Figures 2 to 4 show the association of BMI with risk of any PTB, SPTB and MPTB in each dataset obtained from the optimal fractional polynomial model. On each graph, the mean BMI in that dataset is plotted as a reference line. Across all datasets except CPP, the risk of any PTB increased with lower and higher BMI, with the latter largely driven by a sharp increase in MPTB with increasing BMI from the lowest risk levels. In contrast, the risk of SPTB was higher at lower BMIs but remained low or increased only slightly with higher BMI. Table 1 shows the BMI, with 95% CI, at which the predicted risk for any PTB and MPTB was lowest. It was not possible to calculate this for SPTB in most datasets or in the meta-analysed results because the risk did not vary across most of the BMI distribution. The lowest predicted risk of any PTB was at a BMI slightly below the mean in all datasets except CPP, in which the risk decreased monotonically with increasing BMI, and Bradford, in which the lowest risk was above the mean BMI. The lowest risk of MPTB, where calculable, occurred at BMIs between 20.1 kg/m^2^ and 22.5 kg/m^2^. (Full details of the fractional polynomial models and how we reached the final model are provided in Supplementary Text A.2 and Supplementary Table S9, with results given in Supplementary Table S10.)

**Table 1:** Body mass index (BMI) (95% CI) at which the predicted risk of outcomes was lowest

| Dataset | Mean BMI (kg/m<sup>2</sup>) | Any PTB | MPTB |
| --- | --- | --- | --- |
| CPP | 22.8 | NA<sup>a</sup> | NA<sup>b</sup> |
| Norwegian birth registry | 24.3 | 22.0 (21.1, 23.8) | 20.1 (19.8, 20.6) |
| Danish linked data | 24.4 | 23.1 (22.5, 24.0) | 21.3 (21.0, 21.8) |
| CPRD | 25.7 | 23.6 (22.4, 26.3) | 21.2 (20.7, 22.4) |
| South Australian BEBOLD | 26.6 | 24.4 (22.5, 29.8) | 21.5 (20.8, 22.9) |
| NCHS data (USA) | 26.9 | 22.7 (22.6, 22.8) | 20.8 (20.7, 20.9) |
| SAIL databank | 27.3 | 26.6 (23.1, 51.7) | 22.5 (20.8, 31.4) |
| Bradford | 29.3 | 33.9 (23.2, <sup>c</sup>) | NA<sup>a</sup> |
| Meta-analysed data |  | 24.5 (23.1, 30.3) | 21.3 (20.8, 21.9) |

a. Risk decreased across the whole BMI range
b. Insufficient data
c. Upper limit outside the observed BMI range due to small sample size and resulting high variability in the fractional polynomial terms.

**Figure 2.**
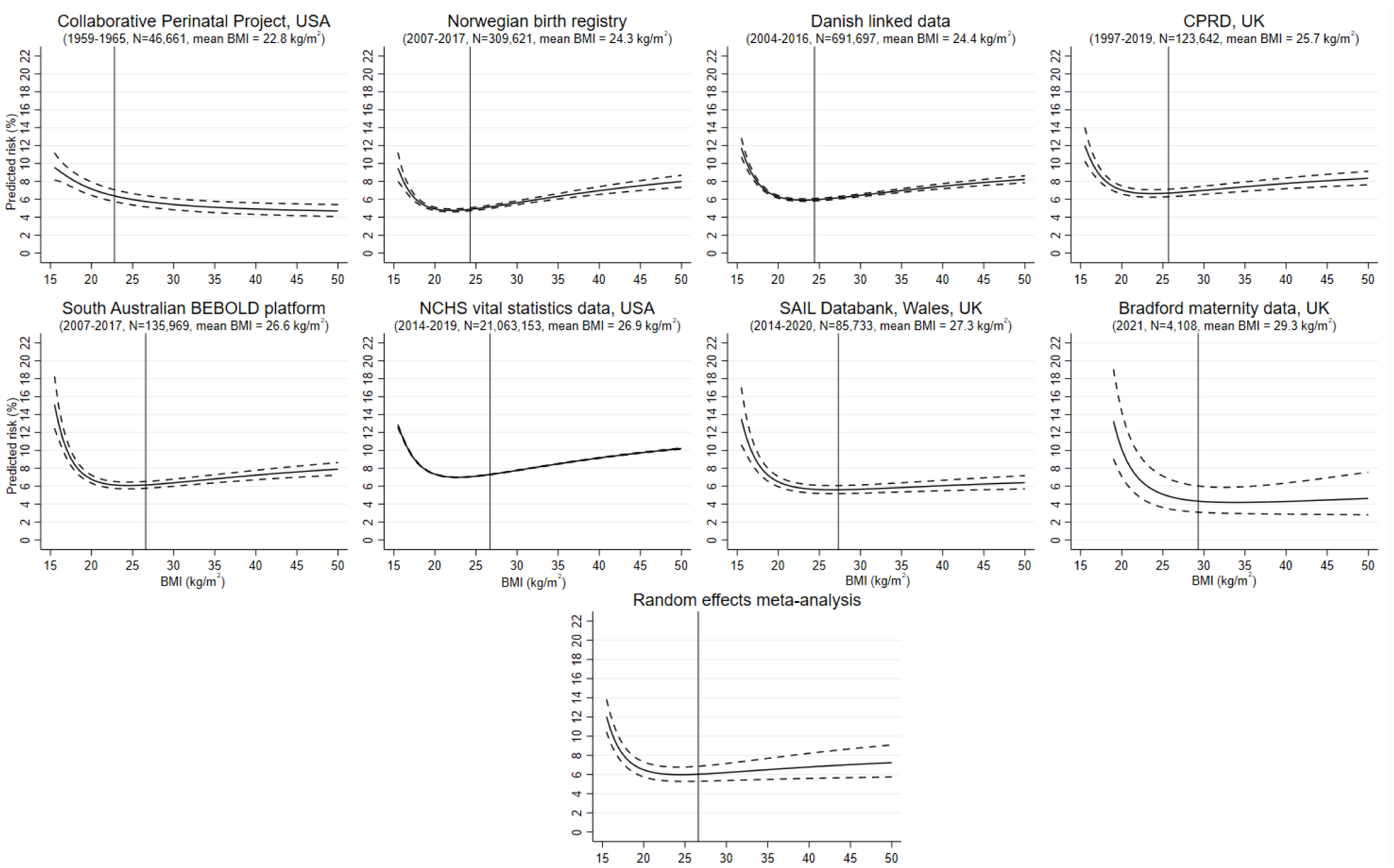
Association of pre-pregnancy BMI with risk of any preterm birth Footnotes: 1. Results are predicted values from the fractional polynomial model with adjustment for covariates, thus represent the predicted risk across the BMI range for individuals in the reference category of all confounders; 2. The vertical reference lines are plotted at the mean BMI for each dataset/overall; 3. CPRD covers the four countries of the UK but we have used a subset linked to HES data, which only covers England.

**Figure 3.**
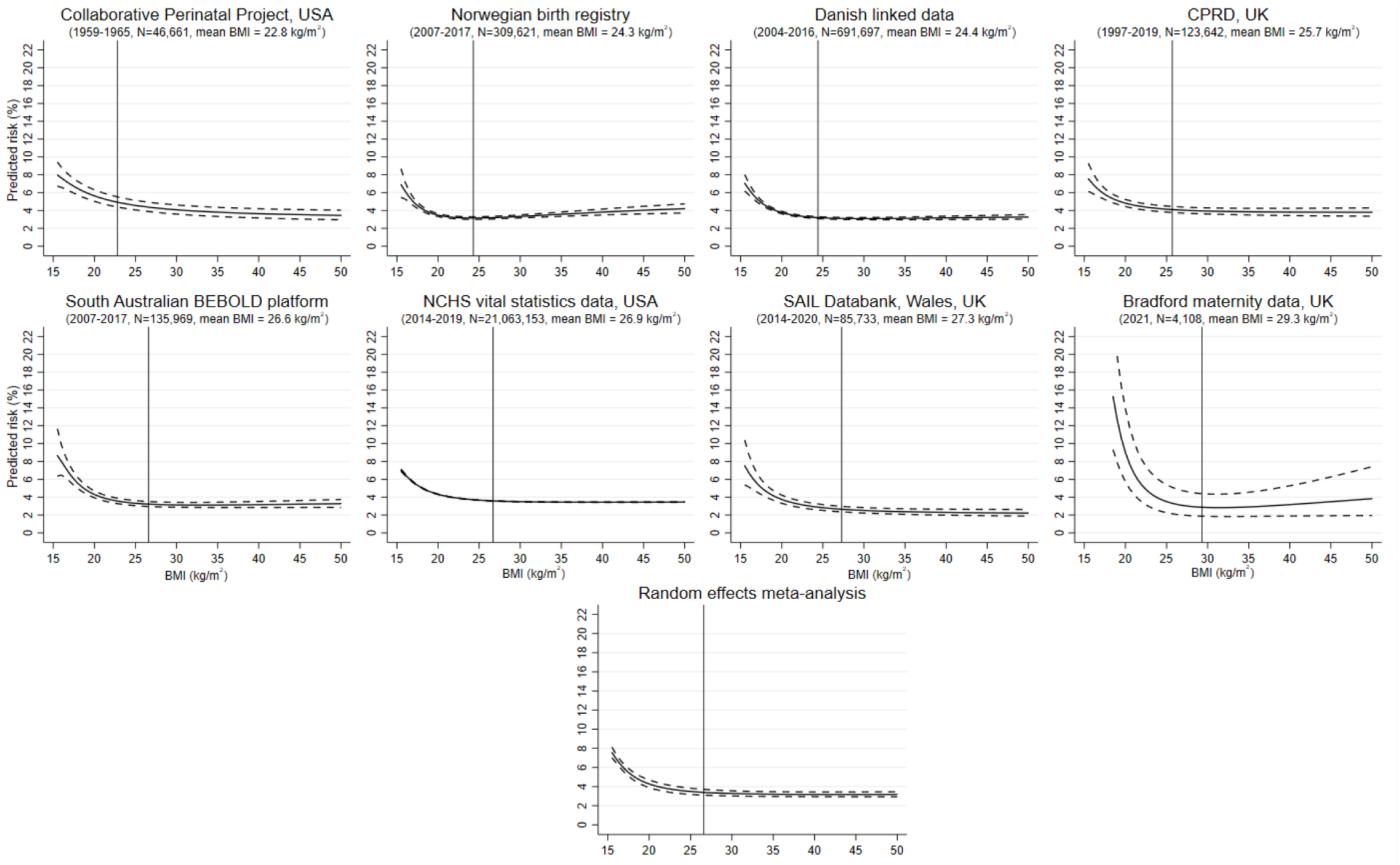
Association of pre-pregnancy BMI with risk of spontaneous preterm birth Footnotes: 1. Results are predicted values from the fractional polynomial model with adjustment for covariates, thus represent the predicted risk across the BMI range for individuals in the reference category of all confounders; 2. The vertical reference lines are plotted at the mean BMI for each dataset/overall; 3. CPRD covers the four countries of the UK but we have used a subset linked to HES data, which only covers England.

**Figure 4.**
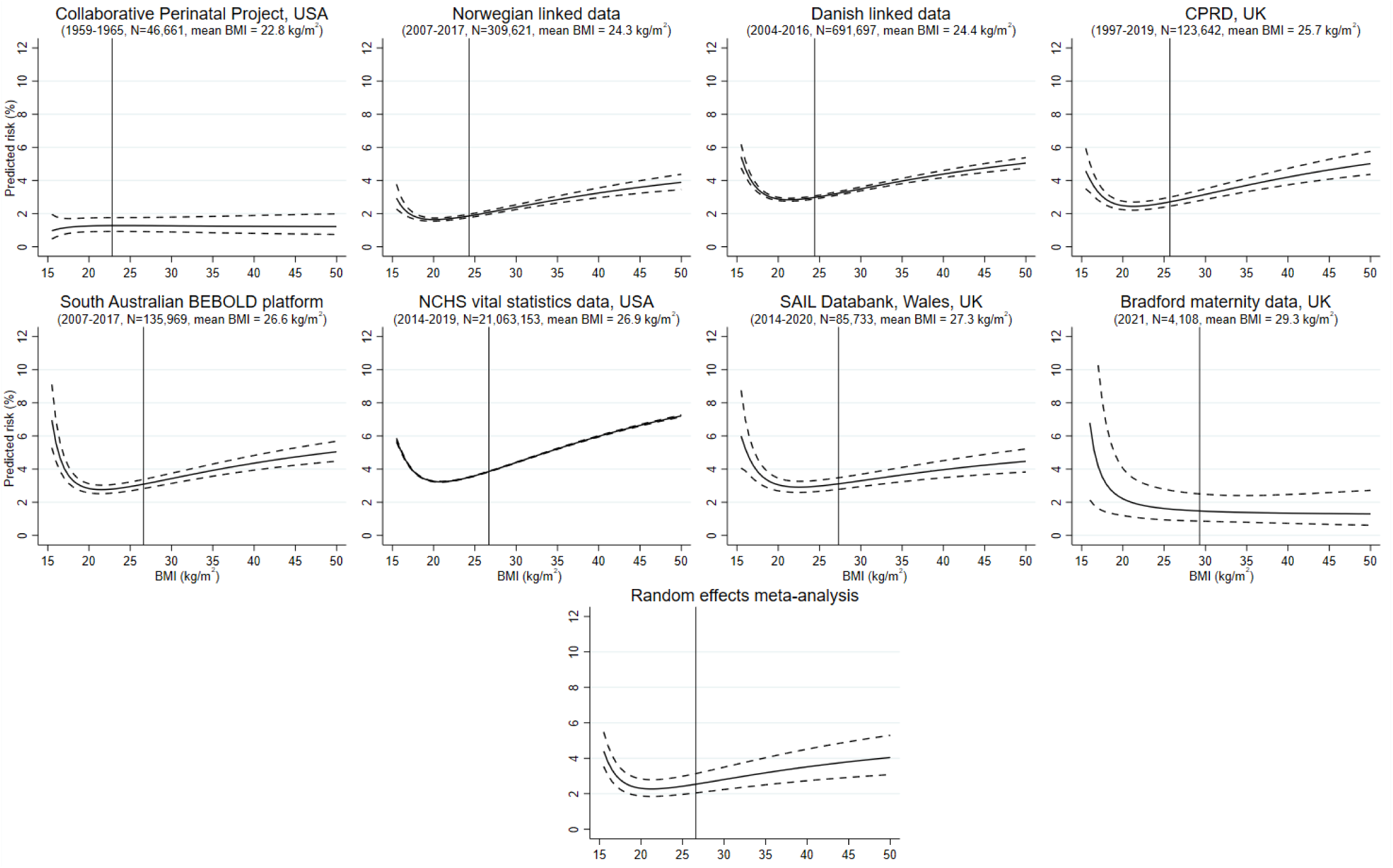
Association of pre-pregnancy BMI with risk of medically indicated preterm birth Footnotes: 1. Results are predicted values from the fractional polynomial model with adjustment for covariates, thus represent the predicted risk across the BMI range for individuals in the reference category of all confounders; 2. The vertical reference lines are plotted at the mean BMI for each dataset/overall; 3. CPRD covers the four countries of the UK but we have used a subset linked to HES data, which only covers England.

### Secondary analyses

The risks of PTB, SPTB and MPTB and adjusted odds ratios using BMI groups are given in Supplementary Tables S11 and S12, respectively. The patterns reflect those shown in Figures 2 to 4. The adjusted odds ratios for very PTB, SPTB and MPTB are given in Supplementary Table S13; these show similar non-linear patterns to those seen for <37 completed weeks.

### Sensitivity analyses

The results from the sensitivity analyses were similar to the overall results (Supplementary Tables S14-S17). Excluding CPP from the meta-analysis made the slope of the curve for any PTB slightly steeper at higher BMIs but had no noticeable impact for SPTB and MTPB (Supplementary Figure S2).

## Discussion

We have examined the relationship between maternal pre-pregnant BMI and any PTB, SPTB, and MPTB in several large datasets from different countries, and have shown non-linear associations with all three outcomes, across all datasets. The higher risk of any PTB at higher BMI was driven by MPTB, whereas the risk of SPTB was increased at lower levels of BMI but remained low or increased only slightly with higher BMI. The key exception to the pattern for any PTB was in the CPP, where a large majority of the preterm births were spontaneous - so the relationship of BMI with any PTB followed that for SPTB, with an increased risk only among underweight women. CPP was based on births between 1959 and 1965, which is around the time that gestational diabetes was first being described and acknowledged^30^.

Similarly, the routine measuring of blood pressure and proteinuria antenatally was not common until the 1960s^31^. Hence MPTB would be expected to be low in this dataset.

In most datasets, the lowest predicted risk of any PTB and MPTB was at a BMI slightly below the mean. In the meta-analysed data, this lowest risk was at a BMI of 24.5 kg/m^2^ for any PTB and 21.3 kg/m^2^ for MPTB. For SPTB, the risk remained relatively constant or increased only slightly for BMIs above 25-30 kg/m^2^. Taken together, these suggest that a healthy BMI to prevent either MPTB or SPTB would be between 21 to 30 kg/m^2^. Apart from the CPP, as already explained, the patterns of association were consistent across the datasets, despite the fact that they reflect different stages of the obesity epidemic, as indicated by the prevalence of overweight and obesity, and had different potential sources of bias due to varying proportions of missing data, measurement error in gestational age and BMI, and possible residual confounding.

Our findings are broadly consistent with previous studies that have explored associations of underweight, overweight, or obesity using conventional BMI categories^4-14^. To our knowledge, two studies have examined the relationship using BMI as a continuous variable. One used locally weighted scatterplot smoothing to examine the association with PTB and found that the minimum risk occurred at a BMI of ∼23.5 kg/m^2 32^. The other applied restricted cubic splines to the US vital statistics data, including births between 2016 and 2018, and found the risk of PTB increased with both low and high BMIs, and was lowest at a BMI of ∼24 kg/m^2 33^. Neither explored associations separately for SPTB and MPTB.

Pre-pregnancy overweight and obesity are both associated with an increased risk of gestational hypertension and gestational diabetes^9,33^, which are associated with increased risk of induction of labour and/or planned caesarean section. This likely explains the increased risk of MPTB with higher BMI. Women who are underweight can have difficulty conceiving and, when they do, are at greater risk of fetal growth restriction and PTB. This may be because of underlying maternal chronic diseases complicating the pregnancy^34^ or maternal undernutrition resulting in impaired fetal growth^35^; these mechanisms likely explain the observed association of lower, but not higher, BMI with SPTB.

The strengths of this work include the inclusion of large datasets from different countries with varying prevalence of obesity. We have used BMI as a continuum to examine non-linear associations and have been able to explore associations with any, MPTB and SPTB. The datasets were generally derived from routine health data, thus minimising the risk of selection bias. That said, selection bias could have arisen due to missing data in some datasets. We undertook complete case analyses as we considered this the least biased approach but acknowledge that there was large variation in the extent of missing data, particularly for BMI. In CPP, gestational age was estimated using LMP, which is less accurate than using early ultrasound measures^36^; further, gestational age was rounded to the nearest week (not completed weeks), which would misclassify some preterm births. In some datasets, weight and height were self-reported, which may be subject to differential measurement error, as overweight women are more likely to underreport their weight^37^.

This would likely mean that the risk would be overestimated for higher BMIs. However, the similarity of non-linear associations across the datasets, despite these variations, suggests that any resulting bias is unlikely to have had a major impact on the general pattern of our findings. To maximise confounder adjustment in each dataset we did not harmonise these to the lowest common denominator. However, residual confounding is possible as some measures were missing or had limited detail in some datasets. For example, in some datasets smoking was categorised as non-smoker/smoker, whereas more detailed measures would provide fuller adjustment. Again, similar results across datasets suggest this has not importantly influenced results. Lastly, we could not identify similar data in low- and middle-income countries (LMIC), where the use of electronic health records for clinical care is still limited and takes priority over their use for research^38^. Thus, our results may not generalise to LMIC populations.

In summary, we have shown a consistent non-linear association between pre-pregnancy BMI and risk of PTB across different populations. Women starting pregnancy with higher BMIs appear to have a higher risk of PTB, but only through medically indicated deliveries. In contrast, underweight women have an increased risk of both SPTB and MPTB. Current antenatal practice identifies and monitors women who may be at risk of MPTB due to pregnancy complications such as gestational diabetes and hypertension. Although the proportion of women who are underweight is relatively small in some of the populations included in this study, in others (e.g. Norway, Denmark) it exceeds the proportion classified as severely obese. Our findings suggest that consideration of the increased risk of SPTB in women with low BMI is also important and that advice to women planning a pregnancy, and clinicians supporting them, should consider both underweight and obesity as risks for PTB.

## Data Availability

CPP and NCHS Vital Statistics data are available online. All proposals to use SAIL Databank data are subject to review by an independent Information Governance Review Panel (IGRP). Once approved, data access is via remote access to a privacy protecting safe haven. Access to CPRD data is subject to approval by the CPRD Research Data Governance (RDG) process. Our CPRD protocol was approved by the Independent Scientific Advisory Committee (ISAC; protocol number 20_145R); approval via ISAC has now been replaced by the RDG process. Perinatal data for the BEBOLD platform are provided by the South Australian Department for Health and Wellbeing. Data are only accessible by researchers who have entered into an agreement with the Data Custodian and are approved by the SA Health Human Research Ethics Committee. The Danish linked data can be made available via remote access to a privacy protecting safe haven at Statistics Denmark after application to the Research Service Center at Statistics Denmark. According to Norwegian legislation, individual-level registry data cannot be made publicly available. The data underlying this project can be accessed by direct application to the Directorate for E-Health pending the required ethical approvals from the Regional Ethical Committees for Medical and Health Research of Norway. Access to NHS data from the Bradford Royal Infirmary is subject to regulations set out by the NHS Health Research Authority.

https://www.archives.gov/research/electronic-records/nih.html

https://www.cdc.gov/nchs/data_access/vitalstatsonline.htm

