## Supplementary materials for "Maternal pre-pregnancy body mass index and risk of preterm birth: a collaboration using large routine health datasets"

A. Supplementary text

Supplementary text A.1: Further details about the cohorts and availability of confounders

Collaborative Perinatal Project (CPP, USA)
This was a multisite, prospective cohort study carried out by the National Institute of Neurological and Communicative Disorders and Stroke. Gestational age at delivery was based on last menstrual period and was recorded in number of weeks, rounded to the nearest week (i.e. not completed weeks). Height was measured and maternal self-reported pre-pregnancy weight was recorded at enrolment (at the first antenatal visit for the majority of women: median; IQR gestational age: 21 weeks; 15-28 weeks); BMI was calculated from these. Maternal age, parity, ethnicity, education, and smoking was also measured at enrolment. At that time there were no ethics review boards but informed consent was obtained from the women.

Norwegian birth registry
The Birth Registry of Norway (MBRN) was established in 1967. Information on maternal self-reported pre-pregnancy height and weight was included in the notification form in the birth registry from 2008 onwards. However, the uptake of this additional registration took some time, resulting in a high proportion of missing in the first years. Estimates of gestational age at delivery was based on routine ultrasound measures taken at 18 gestational weeks, or last menstrual period for the small proportion of deliveries without ultrasound-based estimates (<5%). Maternal age, parity, and smoking was also registered at the time of delivery. No information was available in the Norwegian data on education or ethnicity. The study was approved by the Regional Committee of Medical and Health Research Ethics of South/East Norway. The ethical committee provided a waiver of individual consent for this use of health register data for research in line with Norwegian legislation.

Danish linked data
All individuals living in Denmark are given a unique personal identification number, which can be used to identify individuals in all official registers. This personal identification number was used to link information about individuals from different registers covering the entire Danish population. From the Danish Medical Birth Registry, we identified all births in Denmark between 1 January 2004 and 31 December 2016. The study period started in 2004 as information on pre-pregnancy BMI was not registered until 2004. Information on BMI, gestational age, parity, maternal age, birth interval, smoking, multiple pregnancy, mode of delivery, and characteristics of labour (e.g. whether induced) was obtained from the Medical Birth Registry. Gestational age is based on routine ultrasound measures from early pregnancy or last menstrual period for a small proportion of pregnancies. Maternal pre-pregnancy BMI is calculated based on self-reported weight and height. Educational level and country of origin was obtained from population registers held by Statistics Denmark (https://www.dst.dk/en/TilSalg/Forskningsservice/Data). The processing and linkage of data were approved by the Danish Data Protection Agency (UCHP reference number: 514-0230/18-3000). Ethical approval or informed consent is not required for register-based studies in Denmark.

Clinical Practice Research Datalink (CPRD)
The CPRD GOLD data, including the pregnancy register, are sourced from a sub-sample of primary care centres from across the UK: from those practices that use Vision® GP software (https://cprd.com/sites/default/files/2022-11/2022-11%20CPRD%20GOLD%20Release%20Notes.pdf). These can be linked to a range of other health and health-related datasets. Birth outcome (live or stillborn), delivery details, gestational age at delivery, and maternal age were obtained from the Hospital Episode Statistics (HES) maternity data

(Copyright© 2020, re-used with the permission of NHS Digital. All rights reserved); parity and birth interval from the pregnancy register; and BMI (recorded in the data as BMI itself or derived from recorded height and weight) and smoking from the primary care data. Gestational age in completed weeks is derived from ultrasound or – if not available - last menstrual period. We required BMI to be measured a maximum of 12 months pre-pregnancy and/or a maximum of 15 weeks gestation. CPRD has National Research Ethics Service Committee (NRES) approval for research using the primary care and linked datasets. Individuals registered with participating GP practices are included in the CPRD dataset unless they specifically opt out. The CPRD study protocol was approved by the Independent Scientific Advisory Committee (ISAC; protocol number: 20_145R).

South Australian Better Evidence Better Outcomes Linked Data (BEBOLD) platform
Pregnancy data was obtained from the BEBOLD platform, which includes the South Australian Perinatal Statistics Collection 2007-2016. This is a mandatory collection of all births at least 400 grams or 20 weeks gestation. Maternal height and weight were reported at the first antenatal visit. Over 85% of women attend prior to 14 weeks pregnancy, with height and weight data not reported after 20 weeks gestation. Collection of height and weight data commenced in 2007, with a higher proportion of missing information in the first year of collection. Gestational age was determined from the first day of the last menstrual period if dates were reliable and early ultrasound (up to 20 weeks). A clinical examination could be used in the absence of these data or where there was uncertainty. Approval for the BEBOLD platform was obtained from the South Australian Department of Health's Human Research Ethics Committee, which included a waiver of individual consent for the use of de-identified administrative data.

US National Center for Health Statistics vital statistics (birth registration) data
US states are required to record births and deaths via certificates and Federal law mandates national collection and publication of these data. These data are compiled by the National Center for Health Statistics (NHCS), anonymised and made publicly available; they form the National Vital Statistics System. BMI was included in the fetal death data files from 2014 onwards and, at the time of analysis, data on fetal deaths were available up to the end of 2019. Each record in these datasets relates to a live birth or fetal death, rather than a pregnancy and there are no pregnancy or person-level identifiers in the dataset. To identify pregnancies we matched multiple births occurring close in time in which birth and maternal characteristics were the same. Gestational age at delivery was based on routine ultrasound measurements or last menstrual period for the small proportion (<1%) with no ultrasound-based data. Maternal pre-pregnancy weight and height were self-reported by the women at the time of birth. These publicly available datasets are anonymised.

Welsh linked data: Secure Anonymised Information Linkage (SAIL) Databank
Linkable datasets from the SAIL Databank are made available for approved analyses via a secure research environment, the UK Secure Research Platform (SeRP) [1, 2]. The datasets used for the current study were primary care records, the National Community Child Health (NCCH) Database NCCHD (birth registration plus child health and immunisation data), the Welsh Demographic Service Dataset (demographic characteristics of individuals registered with a GP in Wales) and the Maternity Indicators Dataset (MID) [3] (data - from 2014 onwards - on women from their first antenatal assessment together with data on labour and birth). The MID and the NCCH datasets provided all variables except socio-economic position (Index of Multiple Deprivation (IMD) 2014, which came from the Welsh Demographic Service Dataset, and BMI, which either came from the MID (derived from recorded height and weight) or the primary care data (recorded as BMI itself or derived from recorded height and weight). If BMI came from the primary care data, we required it to be measured a maximum of 12 months pre-pregnancy and/or a maximum of 15 weeks gestation. Gestational age at delivery is based on early ultrasound if available or – if not available – last menstrual period. Details regarding ethics and consent have been described previously [2]; individuals are able to opt out of their data being transferred to SAIL.

Bradford maternity data (UK)
The Bradford Royal Infirmary is a large teaching hospital in Bradford, England, operated by the Bradford Teaching Hospitals NHS Foundation Trust. Gestational age at delivery was based on early ultrasound if available or – if not available – last menstrual period. BMI was derived from height and weight, which were measured by clinical staff at the first antenatal appointment. All data were anonymised and therefore patient consent and ethical approval was not required.

Availability of confounders
Socio-economic position (SEP) was measured differently across the datasets: years of education, index of multiple deprivation (IMD), Townsend score (another area-based deprivation index), and occupation. Ethnicity is not recorded in the Danish or Norwegian registries; in the Danish data, country of origin was included instead. We were able to adjust for either birth or pregnancy interval in all datasets except the Bradford maternity data; where a dataset had both birth and pregnancy interval, we used the most complete. Depending on the dataset, smoking was recorded as either (i) current smoker/non-smoker, (ii) ever/never smoked, or (iii) smoking during pregnancy, with information collected in each trimester (see Supplementary Tables S1-S8). Maternal age (in years) and parity (total number of previous births) were available and categorised in the same way in all datasets (maternal age categorised as: <25, 25-29, 30-34, 35-39, and 40+ years, although presented as mean (SD) years in Supplementary Tables S1-S8; parity categorised as 0, 1, 2, 3, 4+).

Supplementary text A.2: Further details of statistical methods

a) Fractional polynomial models

To fit the fractional polynomials, BMI was first scaled [scaled BMI = (BMI-10)/5]. This is done because if the values of the variable are too large (or too small), this can generate extreme values with certain powers of this variable (e.g. cubic or squared reciprocal powers). Supplementary Table S10 gives the deviance for the different two-power and three-power fractional polynomials for each outcome in each dataset. Model fit was assessed using the change in deviance. Since the three-power models fit better in all datasets except Connected Bradford and CPP and because there was more consistency in terms of the best-fitting three power models, we selected the optimal model from among those with three powers of scaled BMI. For all three outcomes – any preterm birth (PTB), spontaneous preterm birth (SPTB) and medically indicated PTB (MPTB), the optimal model had terms of scaled BMI of -2, -2, -2 (i.e. 1/scaled BMI$^2$, ln(scaled BMI) x 1/scaled BMI$^2$ and ln(scaled BMI) x ln(scaled BMI) x 1/scaled BMI$^2$). For any preterm birth, this polynomial was the best fitting model in three datasets and the second-best fitting in three; for SPTB it was the best fitting model in six of the eight datasets; and for medically indicated PTB it was the best fitting model in three datasets and the second-best in three.

b) Predicted risks and meta-analyses

For each dataset, the fitted (logistic) model was of the form:

$$\begin{aligned}\text{Log odds (PTB)} = \propto + (\beta_1 \times \text{sBMI}^{-2}) + \left(\beta_2 \times \ln(\text{sBMI}) \times \text{sBMI}^{-2}\right) \\ + \left(\beta_3 \times \ln(\text{sBMI}) \times \ln(\text{sBMI}) \times \text{sBMI}^{-2}\right) + \boldsymbol{X}\gamma\end{aligned} \quad [1]$$

where sBMI is scaled BMI, $\mathbf{X}$ represents the confounders, and $\gamma$ the vector of coefficients for the confounders obtained from the logistic regression and with equivalent models for SPTB and MPTB. The study-specific estimates of the constant, α, and fractional polynomial terms, $\beta_1, \beta_2, \beta_3$, were used to plot the predicted risk of PTB against BMI for individuals in the reference category of all confounders for each study.

We used multivariate random effects meta-analysis to pool the fractional polynomial terms (i.e. the $\beta$s, the regression coefficients for the powers of sBMI) and the constant term, $\alpha$, and then used the pooled estimates to plot the predicted (pooled) risk of PTB against BMI.

c) Obtaining the estimated BMI at which the risk of PTB is lowest

These were obtained by differentiating the function given in equation [1] with respect to (scaled) BMI – minimum points occur where the value of this differential is equal to zero.

To calculate confidence intervals, the estimates of α, $\beta_1, \beta_2, \text{and } \beta_3$, together with their variance covariance matrix were used to generate 100,000 bootstrapped samples of these coefficients using the drawnorm function in Stata and the value of BMI at which the minimum point occurred was calculated for each sample. The confidence limits were obtained by taking the 2.5$^{th}$ and 97.5$^{th}$ percentiles of these.

### B. Supplementary tables and figures

Supplementary Figure S1: Summary of missing data

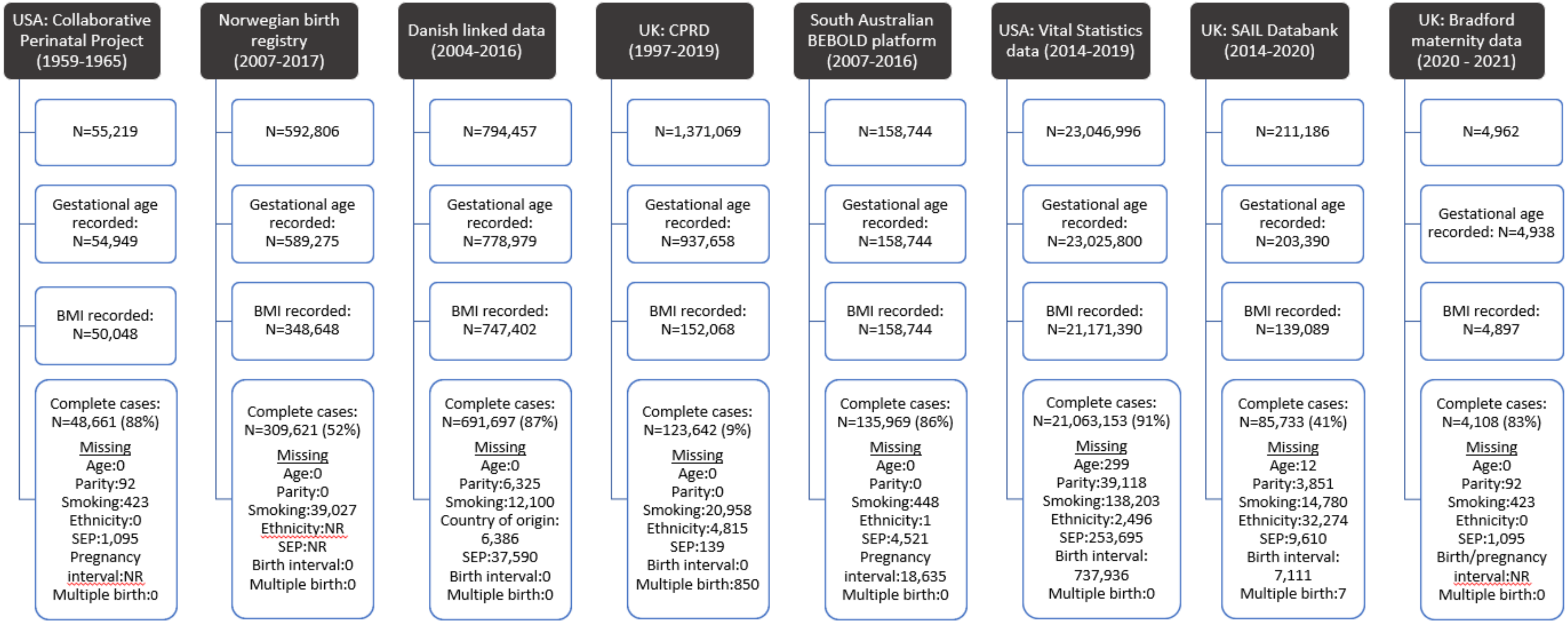

Supplementary Table S1: Characteristics of the whole sample compared to complete cases: Collaborative Perinatal Project (USA, 1959-1965)

| Characteristic | | Whole sample[1] | Complete cases (N=48,661) |
|---|---|---|---|
| | | Frequency (%) | Frequency (%) |
| Mother's age (years) | Mean (SD) | 24 (6.0) | 24 (6.0) |
| Parity | 0 | 16,449 (29.9%) | 14,642 (30.1%) |
| | 1 | 12,571 (22.8%) | 11,108 (22.8%) |
| | 2 | 9,021 (16.4%) | 7,943 (16.3%) |
| | 3 | 6,203 (11.3%) | 5,456 (11.2%) |
| | 4+ | 10,793 (19.6%) | 9,509 (19.5%) |
| Smoking status | Non-smoker | 29,084 (53.3%) | 26,071 (53.6%) |
| | Smoker | 25,531 (46.8%) | 22,587 (46.4%) |
| Ethnicity | White | 25,344 (45.9%) | 21,249 (43.7%) |
| | Black | 25,778 (46.7%) | 23,831 (50.0%) |
| | Other[2] | 4,097 (7.4%) | 3,578 (7.4%) |
| SEP: Maternal education | < High school | 31,138 (57.8%) | 28,411 (58.4%) |
| | High school | 16,320 (30.3%) | 14,701 (30.2%) |
| | > High school | 6,374 (11.8%) | 5,546 (11.4%) |
| Pregnancy size | Singleton | 54,594 (98.9%) | 48,119 (98.9%) |
| | Multiple | 613 (1.1%) | 549 (1.1%) |
| BMI (kg/m$^2$) | Mean (SD) | 22.8 (4.3%) | 22.8 (4.3%) |
| BMI (kg/m$^2$) | <18.5 | 4,629 (9.2%) | 4,479 (9.2%) |
| | 18.5-24.9 | 34,426 (68.5%) | 33,325 (68.5%) |
| | 25-29.9 | 7,829 (15.6%) | 7,594 (15.6%) |
| | 30-34.9 | 2,375 (4.7%) | 2,296 (4.7%) |
| | 35+ | 1,011 (2.0%) | 964 (2.0%) |
| Preterm | Yes | 8,552 (15.6%) | 7,365 (15.1%) |

1. Denominators vary because not all variables are complete

Supplementary Table S2: Characteristics of the whole sample compared to complete cases: Norwegian birth registry (2008-2017)

| Characteristic | | Whole sample[1] | Complete cases (N=309,621) |
|---|---|---|---|
| | | Frequency (%) | Frequency (%) |
| Mother's age (years) | Mean (SD) | 30 (5.2) | 30 (5.1) |
| Parity | 0 | 250,125 (42.5%) | 134,726 (43.5%) |
| | 1 | 214,078 (36.3%) | 111,387 (36.0%) |
| | 2 | 89,200 (15.1%) | 45,882 (14.8%) |
| | 3 | 24,057 (4.1%) | 12,083 (3.9%) |
| | 4+ | 11,815 (2.0%) | 5,543 (1.8%) |
| Smoking status | Never smoked | 418,173 (84.2%) | 260,838 (84.2%) |
| | Smoked before pregnancy | 37,352 (7.5%) | 25,494 (8.2%) |
| | Stopped early pregnancy | 14,326 (2.9%) | 7,868 (2.5%) |
| | Smoked throughout pregnancy | 27,071 (5.5%) | 15,421 (5.0%) |
| Birth interval | No previous birth | 279,668 (47.5%) | 149,851 (48.4%) |
| | <12 months | 40,044 (6.8%) | 19,642 (6.3%) |
| | 12-23 months | 92,323 (15.7%) | 47,210 (15.3%) |
| | 24+ months | 177,240 (30.1%) | 92,918 (30.0%) |
| Pregnancy size | Singleton | 579,544 (98.4%) | 304,443 (98.3%) |
| | Multiple | 9,731 (1.7%) | 5,178 (1.7%) |
| BMI (kg/m$^2$) | Mean (SD) | 24.3 (4.7) | 24.3 (4.7) |
| BMI (kg/m$^2$) | <18.5 | 14,424 (4.1%) | 12,718 (4.1%) |
| | 18.5-24.9 | 215,602 (61.8%) | 191,256 (61.8%) |
| | 25-29.9 | 77,289 (22.2%) | 68,679 (22.2%) |
| | 30-34.9 | 28,909 (8.3%) | 25,815 (8.3%) |
| | 35-39.9 | 9,520 (2.7%) | 8,546 (2.8%) |
| | 40+ | 2,904 (0.8%) | 2,607 (0.8%) |
| Preterm | Yes | 33,358 (5.7%) | 16,689 (5.4%) |

1. Denominators vary because not all variables are complete

Supplementary Table S3: Characteristics of the whole sample compared to complete cases: Danish linked data (2004-2016)

| Characteristic | | Whole sample[1] | Complete cases (N=691,697) |
|---|---|---|---|
| | | Frequency (%) | Frequency (%) |
| **Danish linked data (Denmark)** | | | |
| Mother's age (years) | Mean (SD) | 30 (5.0) | 30 (4.9) |
| Parity | 0 | 352,274 (45.7%) | 314,153 (45.4%) |
| | 1 | 281,338 (36.5%) | 255,471 (36.9%) |
| | 2 | 102,350 (13.3%) | 92,073 (13.3%) |
| | 3 | 24,552 (3.2%) | 21,219 (3.1%) |
| | 4+ | 11,130 (1.4%) | 8,781 (1.3%) |
| Smoking status | Never | 663,270 (86.7%) | 599,267 (86.6%) |
| | Stopped 1st trimester | 17,665 (2.3%) | 16,267 (2.4%) |
| | Smoker | 84,458 (11.0%) | 76,163 (11.0%) |
| Country of origin | Denmark | 657,321 (83.9%) | 603,705 (87.3%) |
| | Other western country | 34,689 (4.4%) | 23,482 (3.4%) |
| | Non-western country | 91,126 (11.6%) | 64,510 (9.3%) |
| Birth interval | No previous birth | 370,425 (46.6%) | 310,909 (45.0%) |
| | <12 months | 2,867 (0.4%) | 2,177 (0.3%) |
| | 12-23 months | 73,555 (9.3%) | 64,651 (9.4%) |
| | 24+ months | 347,610 (43.8%) | 313,960 (45.4%) |
| SEP: Maternal education | < Secondary | 136,685 (18.3%) | 123,710 (17.9%) |
| | Secondary/post-secondary/short-cycle tertiary | 328,645 (43.9%) | 303,661 (43.9%) |
| | Bachelor's degree/higher | 282,894 (37.8%) | 264,326 (38.2%) |
| Pregnancy size | Singleton | 776,567 (97.8%) | 675,941 (97.7%) |
| | Multiple | 17,890 (2.3%) | 15,756 (2.3%) |
| BMI ($kg/m^2$) | Mean (SD) | 24.3 (5.0) | 24.4 (5.0) |
| BMI ($kg/m^2$) | <18.5 | 32,356 (4.3%) | 28,858 (4.2%) |
| | 18.5-24.9 | 465,893 (62.3%) | 430,304 (62.2%) |
| | 25-29.9 | 157,523 (21.1%) | 146,361 (21.2%) |
| | 30-34.9 | 60,316 (8.1%) | 56,441 (8.2%) |
| | 35-39.9 | 21,488 (2.9%) | 20,253 (2.9%) |
| | 40+ | 10,040 (1.3%) | 9,480 (1.4%) |
| Preterm | Yes | 47,531 (6.1%) | 40,748 (5.9%) |

1. Denominators vary because not all variables are complete

Supplementary Table S4: Characteristics of the whole sample compared to complete cases: Clinical Practice Research Datalink (UK, 1997-2019)

| Characteristic | | Whole sample[1] | Complete cases (N=123,642) |
|---|---|---|---|
| | | Frequency (%) | Frequency (%) |
| Mother's age (years) | Mean (SD) | 29 (6.0) | 30 (5.8) |
| Parity | 0 | 689,861 (50.3%) | 62,871 (50.9%) |
| | 1 | 449,437 (32.8%) | 43,882 (35.5%) |
| | 2 | 156,473 (11.4%) | 12,359 (10.0%) |
| | 3 | 49,983 (3.7%) | 3,328 (2.7%) |
| | 4+ | 25,315 (1.9%) | 1,202 (1.0%) |
| Smoking status | Never smoked | 460,522 (46.6%) | 57,865 (46.8%) |
| | Current/ex-smoker | 527,956 (53.4%) | 65,777 (53.2%) |
| Ethnicity | White | 1,133,171 (84.8%) | 103,975 (84.1%) |
| | Black | 60,732 (4.6%) | 5,299 (4.3%) |
| | Asian | 91,352 (6.8%) | 8,895 (7.2%) |
| | Mixed/other | 50,398 (3.8%) | 5,473 (4.4%) |
| Birth interval | No previous birth | 689,590 (50.3%) | 62,830 (50.8%) |
| | <12 months | 21,596 (1.6%) | 1,198 (1.0%) |
| | 12-23 months | 162,209 (11.8%) | 13,976 (11.3%) |
| | 24+ months | 497,674 (36.3%) | 45,638 (36.9%) |
| SEP: Townsend quintile | Least deprived | 245,500 (17.9%) | 22,268 (18.0%) |
| | 2 | 248,903 (18.2%) | 22,265 (18.0%) |
| | 3 | 278,498 (20.3%) | 25,172 (20.4%) |
| | 4 | 316,351 (23.1%) | 28,862 (23.3%) |
| | Most deprived | 280,066 (20.5%) | 25,075 (20.3%) |
| Pregnancy size | Singleton | 1,139,837 (97.3%) | 120,602 (97.5%) |
| | Multiple | 32,227 (2.8%) | 3,040 (2.5%) |
| BMI (kg/m$^2$) | Mean (SD) | 25.6 (5.8) | 25.7 (5.8) |
| BMI (kg/m$^2$) | <18.5 | 9,147 (4.3%) | 5,237 (4.2%) |
| | 18.5-24.9 | 111,248 (52.3%) | 63,774 (51.6%) |
| | 25-29.9 | 52,829 (24.8%) | 30,750 (24.9%) |
| | 30-34.9 | 24,022 (11.3%) | 14,438 (11.7%) |
| | 35-39.9 | 10,082 (4.7%) | 6,222 (5.0%) |
| | 40+ | 5,330 (2.5%) | 3,221 (2.6%) |
| Preterm | Yes | 76,882 (8.2%) | 10,760 (8.7%) |

1. Denominators vary because not all variables are complete

Supplementary Table S5: Characteristics of the whole sample compared to complete cases: South Australian BEBOLD platform (2007-2016)

| Characteristic | | Whole sample[1] | Complete cases (N=158,744) |
|---|---|---|---|
| | | Frequency (%) | Frequency (%) |
| Mother's age (years) | Mean (SD) | 30 (6.0) | 29 (5.5) |
| Parity | 0 | 67,835 (42.7%) | 56,932 (41.9%) |
| | 1 | 54,911 (34.6%) | 48,216 (35.5%) |
| | 2 | 22,669 (14.3%) | 19,529 (14.4%) |
| | 3 | 7,981 (5.0%) | 6,785 (5.0%) |
| | 4+ | 5,348 (3.4%) | 4,507 (3.3%) |
| Smoking status | Non smoker | 131,639 (83.2%) | 114,474 (84.2%) |
| | Quit before 1st antenatal appt | 5,904 (3.7%) | 4,751 (3.5%) |
| | Current smoker | 20,753 (13.1%) | 16,744 (12.3%) |
| Ethnicity | Caucasian | 125,500 (79.1%) | 108,789 (80.0%) |
| | Indigenous | 5,150 (3.2%) | 4,169 (3.1%) |
| | Asian | 18,916 (11.9%) | 15,429 (11.4%) |
| | Other | 9,177 (5.8%) | 7,582 (5.6%) |
| Pregnancy interval | No previous pregnancy | 48,893 (34.9%) | 46,980 (34.6%) |
| | <12 months | 7,388 (5.3%) | 7,201 (5.3%) |
| | 12-23 months | 29,032 (20.7%) | 28,307 (20.8%) |
| | 24+ months | 54,796 (39.1%) | 53,481 (39.3%) |
| SEP: Occupation | Manager, professional, administrator | 34,332 (22.3%) | 30,964 (22.8%) |
| | Para-professional, tradesperson, clerk, sales | 58,621 (38.0%) | 51,698 (38.0%) |
| | Driver, labourer, plant/machine operator | 5,415 (3.5%) | 4,594 (3.4%) |
| | Student, retired, unemployed, home duties | 55,855 (36.2%) | 48,713 (35.8%) |
| Pregnancy size | Singleton | 154,542 (97.4%) | 132,425 (97.4%) |
| | Multiple | 4,202 (2.7%) | 3,544 (2.6%) |
| BMI ($kg/m^2$) | Mean (SD) | 26.6 (6.2) | 26.6 (6.2) |
| BMI ($kg/m^2$) | <18.5 | 4,454 (2.8%) | 3,801 (2.8%) |
| | 18.5-24.9 | 72,220 (45.5%) | 61,941 (45.6%) |
| | 25-29.9 | 43,633 (27.5%) | 37,351 (27.5%) |
| | 30-34.9 | 22,039 (13.9%) | 18,858 (13.9%) |
| | 35-39.9 | 9,999 (6.3%) | 8,582 (6.3%) |
| | 40+ | 6,399 (4.0%) | 5,436 (4.0%) |
| Preterm | Yes | 13,003 (8.2%) | 10,832 (8.0%) |

1. Denominators vary because not all variables are complete

Supplementary Table S6: Characteristics of the whole sample compared to complete cases: National Center for Health Statistics Vital Statistics Data (USA, 2014-2019)

| Characteristic | | Whole sample[1] | Complete cases (N=21,063,153) |
|---|---|---|---|
| | | Frequency (%) | Frequency (%) |
| Mother's age (years) | Mean (SD) | 29 (6) | 29 (6) |
| Parity | 0 | 8,923,569 (38.8%) | 8,460,598 (40.2%) |
| | 1 | 7,330,474 (31.9%) | 6,601,557 (31.3%) |
| | 2 | 3,882,139 (16.9%) | 3,479,600 (16.5%) |
| | 3 | 1,668,185 (7.3%) | 1,486,601 (7.1%) |
| | 4+ | 1,182,410 (5.1%) | 1,034,797 (4.9%) |
| Smoking status | Non-smoker | 20,518,276 (90.6%) | 19,074,067 (90.6%) |
| | Stopped early pregnancy | 750,427 (3.3%) | 712,446 (3.4%) |
| | Smoked through pregnancy | 1,379,343 (6.1%) | 1,276,640 (6.1%) |
| Ethnicity | White | 16,912,884 (73.8%) | 15,674,594 (74.4%) |
| | Black | 3,600,742 (15.7%) | 3,225,160 (15.3%) |
| | Native American/Alaskan | 222,968 (1.0%) | 203,796 (1.0%) |
| | Asian | 1,526,570 (6.7%) | 1,371,605 (6.5%) |
| | Native Hawaiian/Pacific Islander | 74,188 (0.3%) | 60,713 (0.3%) |
| | Mixed race | 572,381 (2.5%) | 527,285 (2.5%) |
| (Live) birth interval | No previous live birth | 8,925,163 (40.4%) | 8,460,598 (40.2%) |
| | <12 months | 190,299 (0.9%) | 179,538 (0.9%) |
| | 12-23 months | 2,793,482 (12.6%) | 2,666,595 (12.7%) |
| | 24+ months | 10,200,823 (46.1%) | 9,756,422 (46.3%) |
| SEP: Maternal education | < High school | 3,088,881 (13.7%) | 2,820,291 (13.4%) |
| | High school | 5,758,080 (25.6%) | 5,359,256 (25.4%) |
| | College, no degree | 6,505,543 (28.9%) | 6,117,492 (29.0%) |
| | Degree/higher | 7,180,330 (31.9%) | 6,755,115 (32.1%) |
| Pregnancy size | Singleton | 22,643,840 (98.3) | 20,719,010 (98.4%) |
| | Multiple | 492,515 (1.7%) | 344,083 (1.6%) |
| BMI (kg/m$^2$) | Mean (SD) | 26.9 (6.7) | 26.9 (6.7) |
| BMI (kg/m$^2$) | <18.5 | 761,335 (3.4%) | 722,846 (3.4%) |
| | 18.5-24.9 | 9,886,682 (44.6%) | 9,400,357 (44.6%) |
| | 25-29.9 | 5,771,361 (26.0%) | 5,468,180 (26.0%) |
| | 30-34.9 | 3,076,717 (13.9%) | 2,915,097 (13.8%) |
| | 35-39.9 | 1,575,300 (7.1%) | 1,495,729 (7.1%) |
| | 40+ | 1,116,104 (5.0%) | 1,060,944 (5.0%) |
| Preterm | Yes | 2,112,879 (9.2%) | 1,869,096 (8.9%) |

1. Denominators vary because not all variables are complete

Supplementary Table S7: Characteristics of the whole sample compared to complete cases: Secure Anonymised Information Linkage (SAIL) Databank (Wales, UK, 2014-2020)

| Characteristic | | Whole sample[1] | Complete cases (N=85,733) |
|---|---|---|---|
| | | Frequency (%) | Frequency (%) |
| Mother's age (years) | Mean (SD) | 29 (5.7) | 29 (5.7) |
| Parity | 0 | 76,108 (39.7%) | 33,590 (39.2%) |
| | 1 | 70,109 (36.5%) | 29,998 (35.0%) |
| | 2 | 30,278 (15.8%) | 14,774 (17.2%) |
| | 3 | 9,795 (5.1%) | 4,831 (5.6%) |
| | 4+ | 5,574 (2.9%) | 2,540 (3.0%) |
| Smoking status | Non-smoker | 123,533 (78.5%) | 67,143 (78.3%) |
| | Smoker | 33,849 (21.5%) | 18,590 (21.7%) |
| Ethnicity | White | 140,252 (88.2%) | 77,304 (90.2%) |
| | Black | 4,812 (3.0%) | 1,768 (2.1%) |
| | Asian | 5,859 (3.7%) | 2,725 (3.2%) |
| | Mixed race | 8,103 (5.1%) | 3,936 (4.6%) |
| Birth interval | No previous birth | 76,108 (39.4%) | 33,590 (39.2%) |
| | <12 months | 2,169 (1.1%) | 838 (1.0%) |
| | 12-23 months | 21,948 (11.4%) | 9,868 (11.5%) |
| | 24+ months | 92,572 (48.1%) | 41,437 (48.3%) |
| IMD decile | Least deprived | 14,606 (7.8%) | 7,064 (8.2%) |
| | 2 | 14,745 (7.9%) | 6,573 (7.7%) |
| | 3 | 15,609 (8.3%) | 6,733 (7.9%) |
| | 4 | 15,560 (8.3%) | 6,409 (7.5%) |
| | 5 | 17,945 (9.6%) | 7,717 (9.0%) |
| | 6 | 18,802 (10.0%) | 8,540 (10.0%) |
| | 7 | 19,837 (10.6%) | 9,069 (10.6%) |
| | 8 | 21,063 (11.2%) | 10,271 (12.0%) |
| | 9 | 22,794 (12.2%) | 10,510 (12.3%) |
| | Most deprived | 26,371 (14.1%) | 12,847 (15.0%) |
| Pregnancy size | Singleton | 205,417 (97.9%) | 83,872 (97.8%) |
| | Multiple | 4,467 (2.1%) | 1,861 (2.2%) |
| BMI ($kg/m^2$) | Mean (SD) | 27.1 (6.4) | 27.3 (6.4) |
| BMI ($kg/m^2$) | <18.5 | 3,495 (2.5%) | 1,932 (2.3%) |
| | 18.5-24.9 | 58,727 (42.0%) | 35,041 (40.9%) |
| | 25-29.9 | 39,673 (28.4%) | 24,564 (28.7%) |
| | 30-34.9 | 21,269 (15.2%) | 13,657 (15.9%) |
| | 35-39.9 | 10,276 (7.4%) | 6,601 (7.7%) |
| | 40+ | 6,305 (4.5%) | 3,938 (4.6%) |
| Preterm | Yes | 15,115 (7.4%) | 6,180 (7.2%) |

1. Denominators vary because not all variables are complete

Supplementary Table S8: Characteristics of the whole sample compared to complete cases: Bradford (UK, 2020-2021)

| | | Whole sample[1] | Complete cases (N=4,938) |
|---|---|---|---|
| | | Frequency (%) | Frequency (%) |
| Mother's age (years) | Mean (SD) | 29 (5.6) | 29 (5.6) |
| Parity | 0 | 2,292 (46.2%) | 1,814 (44.2%) |
| | 1 | 1,481 (29.9%) | 1,226 (29.8%) |
| | 2 | 708 (14.3%) | 627 (15.3%) |
| | 3 | 289 (5.8%) | 265 (6.5%) |
| | 4+ | 192 (3.9%) | 176 (4.3%) |
| Smoking status | Non smoker | 3,895 (78.6%) | 3,194 (77.8%) |
| | Smoker | 1,061 (21.4%) | 914 (22.3%) |
| Ethnicity | White | 1,763 (42.1%) | 1,702 (41.4%) |
| | South Asian | 1,930 (46.1%) | 1,916 (46.6%) |
| | Other | 496 (11.8%) | 490 (11.9%) |
| Pregnancy size | Singleton | 4,893 (98.6%) | 4,056 (98.7%) |
| | Multiple | 69 (1.4%) | 52 (1.3%) |
| BMI (kg/m$^2$) | Mean (SD) | 29.3 (6.2) | 29.3 (6.3) |
| BMI (kg/m$^2$) | <18.5 | 77 (1.6%) | 68 (1.7%) |
| | 18.5-24.9 | 1,187 (24.1%) | 999 (24.3%) |
| | 25-29.9 | 1,642 (33.4%) | 1,358 (33.1%) |
| | 30-34.9 | 1,194 (24.3%) | 989 (24.1%) |
| | 35-39.9 | 537 (10.9%) | 452 (11.0%) |
| | 40+ | 284 (5.8%) | 241 (5.9%) |
| Preterm | Yes | 363 (7.4%) | 290 (7.1%) |

1. Denominators vary because not all variables are complete

Supplementary Table S9: Deviance from different fractional polynomials for each dataset

| | Polynomial terms (two-power models) | | | | | Polynomial terms (three-power models) | | | |
|---|---|---|---|---|---|---|---|---|---|
| **Any PTB** | **0.5 0.5** | **0 0.5** | **-1 -0.5** | **-0.5 -0.5** | | **-2 -2 -1** | **-2 -2 -2** | **0.5 0.5 1** | |
| CPP | 39,481 | 39,482 | 39,481* | 39,482 | | 39,479* | 39,479 | 39,479 | |
| Norwegian | **121,275[a]** | 121,279[c] | 121,376 | 121,371 | | 121,233[d] | **121,231[b]** | 121,260 | |
| Danish | 287,684 | 287,661 | 287,643[c] | **287,627[a]** | | 287,569[d] | **287,554[b]** | 287,591 | |
| CPRD | 69,830[c] | **69,827[a]** | 69,853 | 69,843 | | 69,803[d] | **69,799[b]** | 69,821 | |
| South Australian BEBOLD | 67,178 | 67,167 | **67,140[a]** | 67,142[c] | | 67,135 | 67,134[d] | **67,124[b]** | |
| US Vital Statistics | 11,836,519 | 11,835,481 | 11,834,278[c] | **11,833,809[a]** | | **11,832,917[b]** | 11,833,146[d] | 11,833,447 | |
| SAIL Databank | **41,916[a]** | 41,917[c] | 41,940 | 41,936 | | **41,899[b]** | 41,899[d] | 41,914 | |
| Bradford | 1,940[c] | **1,939[a]** | 1,943 | 1,942 | | 1,941 | 1,943 | 1,939* | |
| **SPTB** | **0.5 0.5** | **-2 -2** | **-0.5 -0.5** | **0, 0** | | **-2 -2 -2** | | | |
| CPP | 35,499* | 35,500 | 35,501 | | | 35,498* | | | |
| Norwegian | **81,233[a]** | 81,264 | 81,282 | 81,261[c] | | **81,213[b]** | | | |
| Danish | 155,613 | 155,612 | 155,608[c] | **155,604[a]** | | **155,583[b]** | | | |
| CPRD | 46,809[c] | **46,804[a]** | 46,812 | | | **46,803[b]** | | | |
| South Australian BEBOLD | 40,545 | 40,537[c] | **40,534[a]** | 40,538 | | **40,529[b]** | | | |
| US Vital Statistics | 6,793,598 | 6,792,982[c] | **6,792,904[a]** | | | **6,792,591[b]** | | | |
| SAIL Databank | 23,604[c] | **23,597[a]** | 23,609 | | | **23,597[b]** | | | |
| Bradford | 1,211 | 1,214 | 1,213 | 1,210* | | 1,208* | | | |
| **MPTB** | **0 0.5** | **3 3** | **-1 -1** | **-0.5 -0.5** | | **-2 -2 -2** | **-2 -2 -1** | **1 1 2** | **0.5 0.5 0.5** |
| US CPP | 6,601 | 6,601 | 6,601* | 6,601 | | 6,601 | 6,601* | 6,601 | 6,601 |
| Norwegian | 61,282[c] | **61,274[a]** | 61,336 | 61,315 | | 61,257[d] | 61,260 | **61,256[b]** | 61,279 |
| Danish | 183,565 | 183,565 | 183,553[c] | **183,527[a]** | | **183,472[b]** | 183,487[d] | 183,503 | 183,488 |
| CPRD | **36,185[a]** | 36,187 | 36,217 | 36,195 | | **36,152[b]** | 36,159[s] | 36,170 | 36,174 |
| South Australian BEBOLD | 39,894 | 39,898 | **39,868[a]** | 39,874[c] | | 39,865[d] | 39,866 | 39,868 | **39,855[b]** |
| US Vital Statistics | 7,163,173 | 7,163,674 | 7,162,446[c] | **7,161,505[a]** | | 7,161,187[d] | **7,160,961[b]** | 7,162,238 | 7,161,286 |
| SAIL Databank | **26,107[a]** | 26,111[c] | 26,125 | 26,117 | | **26,097[b]** | 26,098[d] | 26,104 | 26,106 |
| Bradford | **1,126[a]** | 1,133 | 1,127 | 1,127[c] | | 1,127 | 1,127 | 1,126* | 1,126 |

a Best fitting two-power model; b Best fitting three-power model; c Second-best fitting two-power model; d Second-best fitting three-power model; *Best fitting (two/three-power) model among those displayed. The best fitting models in CPP and Connected Bradford were not necessarily the best/second-best fitting in any other dataset. However, the deviance for the models displayed were very similar (or the same to the nearest whole number) to the deviance for the best-fitting model. CPP: Any PTB: 39,479 (two-power), 39,478 (three-power); SPTB: 35,497 (two-power), 35,495 (three-power); medically indicated PTB: 6601 (both three- and two-power). Connected Bradford: Any PTB 1,934 (three-power), SPTB 1210 (two-power) 1206 (three-power) medically indicated PTB 1123 (three-power)

Supplementary Table S10: Random effects meta-analysis: results are the terms (and their standard errors) for each dataset, the pooled result, and the weights (sBMI=scaled BMI)

| | Term | | | | |
|---|---|---|---|---|---|
| Dataset | Constant | $sBMI^{-2}$ | Ln(sBMI) × $sBMI^{-2}$ | Ln(sBMI) × Ln(sBMI) × $sBMI^{-2}$ | Weight |
| **Any PTB** | | | | | |
| US CPP | -3.11 (0.09) | 0.86 (0.09) | 1.48 (0.30) | 0.54 (0.23) | 13.2% |
| Norwegian | -2.01 (0.09) | -0.11 (0.20) | -1.35 (0.48) | -5.69 (0.85) | 13.1% |
| Danish | -2.09 (0.05) | 0.21 (0.07) | -0.78 (0.18) | -4.39 (0.37) | 13.4% |
| CPRD | -2.16 (0.08) | 0.29 (0.11) | -0.56 (0.27) | -3.30 (0.54) | 13.1% |
| South Australian BEBOLD | -2.15 (0.09) | 0.62 (0.20) | -0.60 (0.53) | -4.32 (0.85) | 13.0% |
| US Vital Statistics | -1.81 (0.006) | -0.03 (0.01) | -0.65 (0.03) | -5.02 (0.06) | 13.8% |
| SAIL Databank | -2.48 (0.11) | 0.79 (0.21) | -0.03 (0.57) | -3.15 (0.97) | 12.7% |
| Bradford | -2.59 (0.52) | 2.06 (1.21) | 6.22 (3.43) | -9.84 (5.54) | 7.6% |
| **Pooled** | -2.33 (0.20) | 0.50 (0.19) | -0.53 (0.07) | -3.04 (1.08) | - |
| **$I^2$** | 99% | 97% | 37% | 99% | - |
| **SPTB** | | | | | |
| US CPP | -3.44 (0.10) | 1.04 (0.10) | 1.68 (0.31) | 0.61 (0.24) | 13.4% |
| Norwegian | -2.81 (0.13) | 0.31 (0.29) | -0.15 (0.70) | -4.71 (1.23) | 13.8% |
| Danish | -3.25 (0.07) | 0.77 (0.10) | 0.69 (0.25) | -2.53 (0.52) | 14.3% |
| CPRD | -3.22 (0.10) | 0.81 (0.11) | 0.71 (0.27) | -0.73 (0.52) | 13.5% |
| South Australian BEBOLD | -3.22 (0.14) | 0.90 (0.34) | 1.86 (0.89) | -3.56 (1.40) | 13.3% |
| US Vital Statistics | -3.28 (0.008) | 0.78 (0.01) | 0.88 (0.04) | -1.34 (0.07) | 16.0% |
| SAIL Databank | -3.82 (0.13) | 1.43 (0.19) | 1.66 (0.47) | -0.60 (0.76) | 12.1% |
| Bradford | -2.30 (0.76) | -0.16 (2.26) | 14.40 (6.18) | -20.55 (8.92) | 3.6% |
| **Pooled** | -3.37 (0.06) | 0.95 (0.10) | 1.12 (0.20) | -1.44 (0.49) | - |
| **$I^2$** | 81% | 86% | 77% | 81% | - |
| **MTPB** | | | | | |
| US CPP | -4.43 (0.37) | -0.30 (0.47) | 0.57 (1.28) | 0.36 (2.12) | 8.3% |
| Norwegian | -2.70 (0.12) | -0.62 (0.20) | -3.08 (0.51) | -5.86 (0.95) | 14.0% |
| Danish | -2.52 (0.06) | -0.18 (0.09) | -1.89 (0.22) | -5.22 (0.46) | 15.3% |
| CPRD | -2.41 (0.12) | -0.53 (0.20) | -1.74 (0.54) | -6.90 (1.03) | 13.8% |
| South Australian BEBOLD | -2.51 (0.11) | 0.21 (0.21) | -2.74 (0.54) | -5.03 (0.95) | 13.7% |
| US Vital Statistics | -1.99 (0.008) | -0.72 (0.02) | -2.04 (0.05) | -7.24 (0.08) | 17.1% |
| SAIL Databank | -2.69 (0.15) | 0.08 (0.34) | -1.09 (0.92) | -4.97 (1.45) | 13.0% |
| Bradford | -4.40 (0.72) | 2.72 (1.31) | -0.98 (4.06) | 0.85 (7.23) | 4.7% |
| **Pooled** | -2.76 (0.18) | -0.15 (0.16) | -1.99 (0.14) | -5.04 (0.58) | - |
| **$I^2$** | 97% | 90% | 44% | 83% | - |

Note: between dataset heterogeneity
The $I^2$ statistics are generally high. This likely reflects a number of features: (i) the overall prevalence of PTB varied across datasets, which would result in heterogeneity in the constant term; (ii) the exact nature of the curves in terms of location of the minimum point (if any) and the extent to which risk increased at low and/or high BMIs would result in heterogeneity in the fractional polynomial terms; (iii) since the $I^2$ statistic is the fraction of the overall variance that is due to between study dataset variance, the fact that the within-dataset estimates were mainly very precise as a result of the large sample sizes would impact on the $I^2$ estimates. Bearing these factors in mind and because the relative consistency in terms of the overall shape of the risk curves with BMI, we deemed it appropriate to meta-analyse the results despite this heterogeneity.

Supplementary Table S11: Univariate risk of any preterm, spontaneous preterm and medically indicated preterm birth by BMI category

| BMI category | CPP, USA | Norwegian birth registry | Danish linked data | CPRD, UK | South Australian BEBOLD | US Vital Statistics data | SAIL Databank, UK | Bradford, UK |
|---|---|---|---|---|---|---|---|---|
| **Any PTB** | | | | | | | | |
| <18.5 | 801 (17.9%) | 815 (6.4%) | 2,130 (7.4%) | 570 (10.9%) | 416 (10.9%) | 75,202 (10.4%) | 207 (10.7%) | 16 (23.5%) |
| 18.5-24.9 | 5,032 (15.1%) | 9,609 (5.0%) | 24,190 (5.6%) | 5,333 (8.4%) | 4,665 (7.5%) | 748,970 (8.0%) | 2,491 (7.1%) | 103 (10.3%) |
| 25-29.9 | 1,071 (14.1%) | 3,838 (5.6%) | 8,542 (5.9%) | 2,627 (8.5%) | 2,770 (7.4%) | 468,977 (8.6%) | 1,684 (6.9%) | 73 (5.4%) |
| 30-34.9 | 336 (14.6%) | 1,672 (6.5%) | 3,684 (6.5%) | 1,332 (9.2%) | 1,647 (8.7%) | 285,104 (9.8%) | 957 (7.0%) | 51 (5.2%) |
| 35-39.9 | 125 (13.0%)[1] | 585 (6.9%) | 1,385 (6.8%) | 585 (9.4%) | 836 (9.7%) | 160,973 (10.8%) | 525 (8.0%) | 26 (5.8%) |
| 40+ | | 170 (6.5%) | 705 (7.4%) | 313 (9.7%) | 498 (9.2%) | 129,870 (12.2%) | 316 (8.0%) | 21 (8.7%) |
| All | 7,365 (15.1%) | 16,689 (5.4%) | 40,748 (5.9%) | 10,760 (8.7%) | 10,832 (8.0%) | 1,869,096 (8.9%) | 6,180 (7.2%) | 290 (7.1%) |
| **SPTB** | | | | | | | | |
| <18.5 | 724 (16.5%) | 546 (4.4) | 1,058 (3.8%) | 365 (7.3%) | 250 (6.9%) | 44,275 (6.4%) | 118 (6.4%) | 12 (17.7%) |
| 18.5-24.9 | 4,388 (13.4%) | 5,869 (3.1) | 11,111 (2.7%) | 3,307 (5.4%) | 2,499 (4.2%) | 385,557 (4.3%) | 1,280 (3.8%) | 59 (5.9%) |
| 25-29.9 | 918 (12.3%) | 2,092 (3.1) | 3,445 (2.4%) | 1,445 (4.9%) | 1,293 (3.6%) | 209,469 (4.0%) | 772 (3.3%) | 34 (2.5%) |
| 30-34.9 | 279 (12.5%) | 862 (3.5) | 1,319 (2.4%) | 630 (4.6%) | 714 (4.0%) | 114,226 (4.2%) | 378 (2.9%) | 22 (2.2%) |
| 35-39.9 | 97 (10.4%)[1] | 278 (3.4) | 456 (2.4%) | 266 (4.5%) | 313 (3.9%) | 57,564 (4.1%) | 193 (3.1%) | 16 (3.5%) |
| 40+ | | 94 (3.7) | 223 (2.5%) | 142 (4.7%) | 193 (3.8%) | 39,636 (4.1%) | 105 (2.8%) | 9 (3.7%) |
| All | 6,406 (13.4%) | 9,741 (3.2%) | 17,612 (2.6%) | 6,155 (5.2%) | 5,263 (4.0%) | 850,727 (4.2%) | 2,846 (3.3%) | 152 (3.7%) |
| **MPTB** | | | | | | | | |
| <18.5 | 51 (1.4%) | 269 (2.2) | 1,072 (3.9%) | 204 (4.2%) | 166 (4.7%) | 29,906 (4.4%) | 88 (4.9%) | ≤ 10[2] |
| 18.5-24.9 | 445 (1.6%) | 3,740 (2.0) | 13,079 (3.1%) | 1,993 (3.3%) | 2,166 (3.6%) | 351,330 (3.9%) | 1,205 (3.6%) | 44 (4.4%) |
| 25-29.9 | 95 (1.4%) | 1,746 (2.6) | 5,209 (3.6%) | 1,165 (4.0%) | 1,477 (4.1%) | 250,955 (4.8%) | 904 (3.8%) | 39 (2.9%) |
| 30-34.9 | 43 (2.2%) | 810 (3.3) | 2,365 (4.3%) | 689 (5.0%) | 934 (5.2%) | 165,468 (5.9%) | 577 (4.4%) | 29 (2.9%) |
| 35-39.9 | 21 (2.4%)[1] | 307 (3.7) | 929 (4.7%) | 317 (5.3%) | 523 (6.3%) | 100,463 (7.0%) | 329 (5.1%) | ≤ 10[2] |
| 40+ | | 76 (3.0) | 482 (5.2%) | 169 (5.5%) | 305 (5.8%) | 87,719 (8.6%) | 210 (5.5%) | 12 (5.0%) |
| All | 655 (1.6%) | 6,948 (2.3%) | 23,136 (3.4%) | 4,537 (3.9%) | 5,571 (4.3%) | 985,841 (4.9%) | 3,313 (3.9%) | 138 (3.4%) |

1. BMI ≥35 kg/m$^2$ as too few individuals with BMI of 40 or higher in this dataset
2. Exact number suppressed for disclosure control purposes

Secondary analyses

Table S12: Adjusted odds ratios (95% CI) of any preterm, spontaneous preterm and medically indicated preterm birth by BMI category

| BMI category | CPP, USA | Norwegian birth registry | Danish linked data | CPRD, UK | South Australian BEBOLD | US Vital Statistics data[2] | SAIL Databank, UK | Bradford, UK |
|---|---|---|---|---|---|---|---|---|
| **Any PTB** | | | | | | | | |
| <18.5 | 1.23 (1.13, 1.34) | 1.33 (1.23, 1.43) | 1.32 (1.26, 1.39) | 1.34 (1.22, 1.48) | 1.54 (1.38, 1.73) | 1.318 (1.307, 1.329) | 1.50 (1.29, 1.75) | 3.01 (1.64, 5.42) |
| 18.5-24.9 | 1.00 | 1.00 | 1.00 | 1.00 | 1.00 | 1.00 | 1.00 | 1.00 |
| 25-29.9 | 0.83 (0.77, 0.89) | 1.11 (1.07, 1.16) | 1.02 (1.00, 1.05) | 1.01 (0.96, 1.06) | 0.95 (0.90, 1.00) | 1.030 (1.025, 1.034) | 0.94 (0.88, 1.00) | 0.53 (0.38, 0.72) |
| 30-34.9 | 0.82 (0.73, 0.93) | 1.28 (1.21, 1.36) | 1.11 (1.07, 1.16) | 1.09 (1.02, 1.16) | 1.10 (1.03, 1.17) | 1.148 (1.143, 1.154) | 0.92 (0.85, 0.99) | 0.46 (0.32, 0.67) |
| 35-39.9 | 0.70 (0.57, 0.85)[1] | 1.38 (1.26, 1.52) | 1.18 (1.12, 1.26) | 1.12 (1.02, 1.23) | 1.24 (1.14, 1.34) | 1.249 (1.241, 1.257) | 1.02 (0.92, 1.13) | 0.57 (0.36, 0.90) |
| 40+ | | 1.32 (1.12, 1.55) | 1.26 (1.16, 1.37) | 1.14 (1.00, 1.29) | 1.05 (0.94, 1.17) | 1.402 (1.393, 1.411) | 1.03 (0.90, 1.16) | 0.75 (0.45, 1.26) |
| **SPTB** | | | | | | | | |
| <18.5 | 1.27 (1.16, 1.38) | 1.42 (1.29, 1.56) | 1.38 (1.29, 1.48) | 1.34 (1.19, 1.50) | 1.66 (1.45, 1.91) | 1.406 (1.392, 1.421) | 1.63 (1.33, 1.99) | 3.69 (1.80, 7.19) |
| 18.5-24.9 | 1.00 | 1.00 | 1.00 | 1.00 | 1.00 | 1.00 | 1.00 | 1.00 |
| 25-29.9 | 0.82 (0.76, 0.89) | 1.00 (0.95, 1.06) | 0.90 (0.87, 0.94) | 0.91 (0.85, 0.97) | 0.83 (0.77, 0.89) | 0.897 (0.892, 0.902) | 0.85 (0.77, 0.93) | 0.46 (0.29, 0.69) |
| 30-34.9 | 0.80 (0.70, 0.92) | 1.09 (1.01, 1.17) | 0.89 (0.84, 0.94) | 0.84 (0.77, 0.92) | 0.88 (0.81, 0.96) | 0.891 (0.885, 0.898) | 0.71 (0.63, 0.80) | 0.38 (0.23, 0.63) |
| 35-39.9 | 0.64 (0.51, 0.79)[1] | 1.07 (0.94, 1.21) | 0.86 (0.78, 0.95) | 0.83 (0.73, 0.95) | 0.85 (0.75, 0.96) | 0.863 (0.856, 0.871) | 0.74 (0.64, 0.87) | 0.66 (0.37, 1.17) |
| 40+ | | 1.19 (0.96, 1.48) | 0.88 (0.77, 1.02) | 0.85 (0.72, 1.02) | 0.75 (0.64, 0.87) | 0.825 (0.816, 0.834) | 0.67 (0.55, 0.83) | 0.58 (0.29, 1.21) |
| **MPTB** | | | | | | | | |
| <18.5 | 0.94 (0.70, 1.27) | 1.16 (1.02, 1.32) | 1.27 (1.19, 1.36) | 1.35 (1.16, 1.57) | 1.40 (1.18, 1.66) | 1.192 (1.177, 1.207) | 1.36 (1.09, 1.71) | 1.65 (0.57, 4.81) |
| 18.5-24.9 | 1.00 | 1.00 | 1.00 | 1.00 | 1.00 | 1.00 | 1.00 | 1.00 |
| 25-29.9 | 0.77 (0.61, 0.97) | 1.32 (1.21, 1.36) | 1.13 (1.09, 1.17) | 1.17 (1.09, 1.27) | 1.10 (1.02, 1.18) | 1.177 (1.171, 1.183) | 1.03 (0.94, 1.12) | 0.68 (0.43, 1.07) |
| 30-34.9 | 1.08 (0.78, 1.49) | 1.58 (1.45, 1.71) | 1.31 (1.24, 1.37) | 1.50 (1.36, 1.64) | 1.38 (1.27, 1.51) | 1.436 (1.427, 1.445) | 1.13 (1.02, 1.25) | 0.63 (0.39, 1.03) |
| 35-39.9 | 1.12 (0.71, 1.76)[1] | 1.85 (1.63, 2.10) | 1.46 (1.35, 1.57) | 1.60 (1.41, 1.82) | 1.73 (1.55, 1.92) | 1.686 (1.673, 1.700) | 1.30 (1.15, 1.48) | 0.52 (0.26, 1.05) |
| 40+ | | 1.52 (1.19, 1.92) | 1.56 (1.41, 1.74) | 1.60 (1.36, 1.90) | 1.42 (1.24, 1.63) | 2.051 (2.035, 2.068) | 1.38 (1.18, 1.61) | 1.04 (0.53, 2.04) |

1. BMI ≥35 $kg/m^2$ as too few individuals with BMI>40 in this dataset
2. These ORs given to 3 decimal places to distinguish limits from point estimate (not possible with 2dp)

Supplementary Table S13: Adjusted odds ratios (95% CI) for any very (<32 completed weeks) preterm, spontaneous very preterm and medically indicated very preterm birth by BMI category

| **Any very preterm birth** | | | | | | | | |
|---|---|---|---|---|---|---|---|---|
| BMI category | CPP, USA | Norwegian birth registry | Danish linked data | CPRD, UK | South Australian BEBOLD | US vital statistics | SAIL Databank, UK | Bradford, UK |
| <18.5 | 1.18 (1.01, 1.38) | 1.33 (1.08, 1.64) | 1.26 (1.13, 1.42) | 1.21 (1.04, 2.22) | 1.43 (1.06, 1.94) | 1.26 (1.23, 1.28) | 1.57 (1.12, 2.22) | 5.62 (2.12, 14.91) |
| 18.5-24.9 | 1.00 | 1.00 | 1.00 | 1.00 | 1.00 | 1.00 | 1.00 | 1.00 |
| 25-29.9 | 0.87 (0.76, 1.00) | 1.26 (1.13, 1.39) | 1.11 (1.05, 1.18) | 1.01 (0.93, 1.10) | 0.93 (0.81, 1.07) | 1.10 (1.09, 1.11) | 0.93 (0.79, 1.09) | 0.55 (0.28, 1.07) |
| 30-34.9 | 0.89 (0.71, 1.11) | 1.60 (1.39, 1.84) | 1.27 (1.17, 1.38) | 1.09 (0.98, 1.22) | 1.17 (0.99, 1.38) | 1.30 (1.28, 1.31) | 1.02 (0.85, 1.23) | 0.48 (0.23, 1.00) |
| 35-39.9 | 0.79 (0.56, 1.12)[1] | 1.45 (1.14, 1.84) | 1.25 (1.09, 1.44) | 1.16 (1.00, 1.35) | 1.31 (1.06, 1.62) | 1.43 (1.41, 1.45) | 0.99 (0.78, 1.27) | 0.74 (0.31, 1.77) |
| 40+ | | 1.87 (1.29, 2.71) | 1.58 (1.32, 1.88) | 1.02 (0.82, 1.26) | 0.98 (0.74, 1.29) | 1.54 (1.52, 1.57) | 1.06 (0.79, 1.43) | 1.12 (0.47, 2.71) |
| **Spontaneous very preterm birth** | | | | | | | | |
| <18.5 | 1.16 (0.98, 1.38) | 1.26 (0.96, 1.66) | 1.27 (1.06, 1.52) | 1.32 (1.10, 1.59) | 1.92 (1.32, 2.80) | 1.32 (1.28, 1.36) | 1.81 (1.18, 2.78) | 7.47 (2.18, 25.61) |
| 18.5-24.9 | 1.00 | 1.00 | 1.00 | 1.00 | 1.00 | 1.0 | 1.00 | 1.00 |
| 25-29.9 | 0.84 (0.72, 0.98) | 1.07 (0.93, 1.23) | 1.02 (0.93, 1.13) | 0.94 (0.84, 1.04) | 0.80 (0.65, 0.98) | 1.01 (1.00, 1.03) | 0.78 (0.62, 0.97) | 0.43 (0.16, 1.19) |
| 30-34.9 | 0.88 (0.69, 1.13) | 1.40 (1.16, 1.69) | 1.06 (0.92, 1.23) | 0.86 (0.74, 0.99) | 1.06 (0.84, 1.34) | 1.15 (1.13, 1.17) | 0.93 (0.72, 1.20) | 0.45 (0.16, 1.33) |
| 35-39.9 | 0.76 (0.51, 1.12)[1] | 1.10 (0.78, 1.54) | 1.15 (0.92, 1.45) | 0.77 (0.62, 0.97) | 1.14 (0.84, 1.56) | 1.26 (1.23, 1.28) | 0.77 (0.53, 1.10) | 0.67 (0.18, 2.46) |
| 40+ | | 1.94 (1.22, 3.09) | 1.40 (1.05, 1.85) | 0.81 (0.60, 1.09) | 0.88 (0.59, 1.30) | 1.28 (1.25, 1.31) | 0.85 (0.55, 1.31) | 0.64 (0.14, 2.94) |
| **Medically indicated very preterm birth** | | | | | | | | |
| <18.5 | 1.41 (0.79, 2.49) | 1.44 (1.06, 1.97) | 1.25 (1.08, 1.45) | 1.00 (0.75, 1.35) | 1.00 (0.61, 1.64) | 1.23 (1.19, 1.26) | 1.26 (0.71, 2.23) | 3.36 (0.70, 16.13) |
| 18.5-24.9 | 1.00 | 1.00 | 1.00 | 1.00 | 1.00 | 1.00 | 1.00 | 1.00 |
| 25-29.9 | 0.87 (0.52, 1.40) | 1.54 (1.32, 1.79) | 1.17 (1.08, 1.26) | 1.13 (0.99, 1.30) | 1.07 (0.88, 1.29) | 1.17 (1.15, 1.18) | 1.11 (0.88, 1.39) | 0.68 (0.28, 1.62) |
| 30-34.9 | 1.05 (0.52, 2.13) | 1.89 (1.55, 2.32) | 1.40 (1.27, 1.55) | 1.50 (1.27, 1.76) | 1.27 (1.02, 1.60) | 1.41 (1.39, 1.43) | 1.13 (0.86, 1.47) | 0.52 (0.19, 1.42) |
| 35-39.9 | 0.76 (0.24, 2.45)[1] | 1.99 (1.43, 2.76) | 1.31 (1.11, 1.55) | 1.88 (1.53, 2.31) | 1.48 (1.11, 1.98) | 1.58 (1.55, 1.61) | 1.25 (0.89, 1.74) | 0.84 (0.26, 2.29) |
| 40+ | | 1.73 (0.95, 3.16) | 1.67 (1.34, 2.07) | 1.39 (1.01, 1.90) | 1.11 (0.76, 1.62) | 1.73 (1.70, 1.76) | 1.32 (0.88, 1.98) | 1.61 (0.54, 4.77) |

1. BMI ≥35 $kg/m^2$ as too few individuals with BMI>40 in this dataset

Sensitivity analyses

Supplementary Table S14: Adjusted odds ratios (95% CI) for any preterm, spontaneous preterm and medically indicated preterm birth by BMI category: inverse probability weighted[1]

| | | | | **SPTB** | | | | |
|---|---|---|---|---|---|---|---|---|
| BMI category | CPP, USA | Norwegian birth registry | Danish linked data | CPRD, UK | South Australian BEBOLD | US vital statistics | SAIL Databank, UK | Bradford, UK |
| <18.5 | 1.27 (1.16, 1.38) | 1.42 (1.29, 1.56) | 1.38 (1.29, 1.48) | 1.34 (1.19, 1.50) | 1.70 (1.48, 1.95) | 1.403 (1.388, 1.418) | 1.63 (1.34, 1.99) | 3.59 (1.80, 7.17) |
| 18.5-24.9 | 1.00 | 1.00 | 1.00 | 1.00 | 1.00 | 1.00 | 1.00 | 1.00 |
| 25-29.9 | 0.82 (0.76, 0.89) | 1.00 (0.95, 1.06) | 0.90 (0.86, 0.94) | 0.90 (0.85, 0.97) | 0.84 (0.77, 0.90) | 0.897 (0.892, 0.902) | 0.85 (0.77, 0.93) | 0.46 (0.30, 0.71) |
| 30-34.9 | 0.81 (0.71, 0.92) | 1.08 (1.00, 1.16) | 0.90 (0.84, 0.95) | 0.85 (0.77, 0.92) | 0.91 (0.83, 1.00) | 0.889 (0.883, 0.896) | 0.72 (0.64, 0.81) | 0.39 (0.24, 0.65) |
| 35-39.9 | 0.63 (0.51, 0.79)[2] | 1.07 (0.33, 1.21) | 0.86 (0.78, 0.95) | 0.85 (0.74, 0.97) | 0.85 (0.73, 0.97) | 0.860 (0.852, 0.868) | 0.75 (0.64, 0.88) | 0.67 (0.38, 1.19) |
| 40+ | | 1.20 (0.39, 1.49) | 0.85 (0.73, 0.99) | 0.86 (0.72, 1.03) | 0.77 (0.65, 0.91) | 0.811 (0.801, 0.820) | 0.65 (0.52, 0.81) | 0.60 (0.30, 1.23) |
| | | | | **MPTB** | | | | |
| <18.5 | 0.92 (0.69, 1.24) | 1.16 (1.02, 1.32) | 1.26 (1.18, 1.35) | 1.34 (1.16, 1.56) | 1.44 (1.22, 1.71) | 1.197 (1.182, 1.212) | 1.39 (1.11, 1.73) | 1.65 (0.58, 4.69) |
| 18.5-24.9 | 1.00 | 1.00 | 1.00 | 1.00 | 1.00 | 1.00 | 1.00 | 1.00 |
| 25-29.9 | 0.77 (0.61, 0.98) | 1.28 (1.21, 1.36) | 1.12 (1.08, 1.16) | 1.17 (1.08, 1.26) | 1.10 (1.02, 1.18) | 1.169 (1.163, 1.175) | 1.03 (0.94, 1.12) | 0.67 (0.43, 1.05) |
| 30-34.9 | 1.12 (0.80, 1.55) | 1.55 (1.43, 1.68) | 1.30 (1.24, 1.37) | 1.48 (1.35, 1.62) | 1.39 (1.27, 1.51) | 1.420 (1.411, 1.429) | 1.12 (1.01, 1.25) | 0.63 (0.39, 1.03) |
| 35-39.9 | 1.13 (0.71, 1.78)[2] | 1.81 (1.60, 2.06) | 1.45 (1.35, 1.56) | 1.59 (1.40, 1.81) | 1.70 (1.52, 1.91) | 1.662 (1.649, 1.675) | 1.30 (1.14, 1.48) | 0.52 (0.25, 1.06) |
| 40+ | | 1.49 (1.17, 1.90) | 1.54 (1.39, 1.72) | 1.59 (1.35, 1.89) | 1.41 (1.22, 1.62) | 2.014 (1.997, 2.031) | 1.36 (1.16, 1.60) | 1.08 (0.56, 2.07) |

1. Weighting SPTB by the inverse of one minus the probability of being a medically indicated preterm birth and weighting medically indicated preterm birth by the inverse of one minus the probability of being a SPTB
2. BMI ≥35 $kg/m^2$ as too few individuals with BMI of 40 or higher in this dataset
3. These ORs given to 3 decimal places to distinguish limits from point estimate (not possible with 2dp)

Supplementary Table S15: Adjusted odds ratios (95% CI) for any preterm, spontaneous preterm and medically indicated preterm birth by BMI category excluding stillbirths

| Any PTB | | | | | | | | |
|---|---|---|---|---|---|---|---|---|
| BMI category | CPP, USA | Norwegian birth registry | Danish linked data | CPRD, UK | South Australian BEBOLD | US vital statistics | SAIL Databank, UK | Bradford, UK |
| <18.5 | 1.22 (1.12, 1.33) | 1.35 (1.24, 1.45) | 1.33 (1.26, 1.40) | 1.35 (1.22, 1.48) | 1.56 (1.39, 1.74) | 1.324 (1.314, 1.335) | 1.48 (1.27, 1.73) | 2.72 (1.44, 5.13) |
| 18.5-24.9 | 1.00 | 1.00 | 1.00 | 1.00 | 1.00 | 1.00 | 1.00 | 1.00 |
| 25-29.9 | 0.80 (0.74, 0.86) | 1.11 (1.07, 1.16) | 1.02 (0.99, 1.04) | 1.01 (0.96, 1.06) | 0.95 (0.90, 1.01) | 1.027 (1.023, 1.031) | 0.91 (0.86, 0.98) | 0.51 (0.37, 0.72) |
| 30-34.9 | 0.81 (0.71, 0.93) | 1.28 (1.20, 1.35) | 1.10 (1.06, 1.14) | 1.09 (1.02, 1.16) | 1.11 (1.04, 1.18) | 1.144 (1.139, 1.150) | 0.90 (0.83, 0.97) | 0.44 (0.30, 0.64) |
| 35-39.9 | 0.67 (0.54, 0.82)[1] | 1.39 (1.26, 1.52) | 1.17 (1.10, 1.24) | 1.13 (1.03, 1.24) | 1.23 (1.13, 1.34) | 1.245 (1.238, 1.253) | 1.00 (0.90, 1.11) | 0.60 (0.38, 0.95) |
| 40+ | | 1.29 (1.09, 1.53) | 1.23 (1.12, 1.34) | 1.14 (1.00, 1.29) | 1.06 (0.95, 1.18) | 1.396 (1.387, 1.405) | 0.99 (0.87, 1.13) | 0.78 (0.47, 1.29) |
| **SPTB** | | | | | | | | |
| <18.5 | 1.26 (1.15, 1.37) | 1.41 (1.30, 1.57) | 1.39 (1.30, 1.49) | 1.33 (1.19, 1.50) | 1.68 (1.47, 1.94) | 1.406 (1.392, 1.421) | 1.63 (1.33, 1.99) | 3.34 (1.64, 6.81) |
| 18.5-24.9 | 1.00 | 1.00 | 1.00 | 1.00 | 1.00 | 1.00 | 1.00 | 1.00 |
| 25-29.9 | 0.81 (0.75, 0.88) | 1.01 (0.95, 1.06) | 0.90 (0.86, 0.93) | 0.90 (0.85, 0.97) | 0.83 (0.77, 0.89) | 0.897 (0.892, 0.902) | 0.84 (0.77, 0.93) | 0.45 (0.29, 0.69) |
| 30-34.9 | 0.80 (0.70, 0.92) | 1.08 (1.00, 1.17) | 0.88 (0.83, 0.94) | 0.84 (0.77, 0.92) | 0.89 (0.81, 0.97) | 0.892 (0.886, 0.898) | 0.71 (0.63, 0.80) | 0.38 (0.23, 0.63) |
| 35-39.9 | 0.64 (0.51, 0.80)[1] | 1.08 (0.95, 1.22) | 0.85 (0.77, 0.94) | 0.84 (0.73, 0.95) | 0.85 (0.75, 0.97) | 0.864 (0.856, 0.872) | 0.75 (0.64, 0.88) | 0.67 (0.38, 1.19) |
| 40+ | | 1.18 (0.95, 1.47) | 0.85 (0.73, 0.99) | 0.85 (0.71, 1.02) | 0.75 (0.64, 0.88) | 0.826 (0.817, 0.835) | 0.65 (0.53, 0.81) | 0.61 (0.30, 1.22) |
| **MPTB** | | | | | | | | |
| <18.5 | 0.92 (0.67, 1.25) | 1.19 (1.05, 1.36) | 1.27 (1.18, 1.36) | 1.37 (1.17, 1.59) | 1.40 (1.17, 1.66) | 1.193 (1.178, 1.208) | 1.31 (1.03, 1.66) | 1.37 (0.41, 4.62) |
| 18.5-24.9 | 1.00 | 1.00 | 1.00 | 1.00 | 1.00 | 1.00 | 1.00 | 1.00 |
| 25-29.9 | 0.76 (0.59, 0.96) | 1.29 (1.21, 1.37) | 1.12 (1.08, 1.16) | 1.18 (1.10, 1.28) | 1.11 (1.03, 1.20) | 1.177 (1.170, 1.183) | 0.99 (0.90, 1.09) | 0.67 (0.41, 1.08) |
| 30-34.9 | 1.00 (0.70, 1.42) | 1.58 (1.45, 1.71) | 1.29 (1.22, 1.35) | 1.51 (1.37, 1.66) | 1.40 (1.29, 1.53) | 1.435 (1.426, 1.443) | 1.10 (0.99, 1.22) | 0.58 (0.34, 0.98) |
| 35-39.9 | 1.03 (0.63, 1.70)[1] | 1.88 (1.65, 2.13) | 1.44 (1.34, 1.56) | 1.64 (1.44, 1.86) | 1.74 (1.56, 1.94) | 1.686 (1.673, 1.699) | 1.27 (1.11, 1.45) | 0.54 (0.26, 1.14) |
| 40+ | | 1.47 (1.15, 1.88) | 1.53 (1.37, 1.71) | 1.62 (1.37, 1.92) | 1.45 (1.27, 1.67) | 2.051 (2.034, 2.068) | 1.33 (1.13, 1.57) | 1.09 (0.55, 2.16) |

1. BMI ≥35 $kg/m^2$ as too few individuals with BMI>40 in this dataset
2. These ORs given to 3 decimal places to distinguish limits from point estimate (not possible with 2dp)

Supplementary Table S16: Adjusted odds ratios (95% CI) for any preterm, spontaneous preterm and medically indicated preterm birth by BMI category excluding post term births

| BMI category | CPP, USA | Norwegian birth registry | Danish linked data | CPRD, UK | South Australian BEBOLD | US vital statistics | SAIL Databank, UK | Bradford, UK |
|---|---|---|---|---|---|---|---|---|
| **Any PTB** | | | | | | | | |
| <18.5 | 1.21 (1.11, 1.32) | 1.30 (1.21, 1.41) | 1.30 (1.24, 1.37) | 1.33 (1.21, 1.47) | 1.43 (1.06, 1.94) | 1.317 (1.306, 1.328) | 1.50 (1.28, 1.75) | 3.00 (1.64, 5.51) |
| 18.5-24.9 | 1.00 | 1.00 | 1.00 | 1.00 | 1.00 | 1.00 | 1.00 | 1.00 |
| 25-29.9 | 0.85 (0.79, 0.92) | 1.13 (1.08, 1.17) | 1.04 (1.01, 1.06) | 1.01 (0.96, 1.07) | 0.93 (0.81, 1.07) | 1.029 (1.025, 1.033) | 0.94 (0.88, 1.01) | 0.53 (0.38, 0.73) |
| 30-34.9 | 0.87 (0.76, 0.98) | 1.29 (1.22, 1.37) | 1.13 (1.08, 1.17) | 1.10 (1.03, 1.17) | 1.17 (0.99, 1.38) | 1.147 (1.142, 1.153) | 0.93 (0.86, 1.00) | 0.47 (0.33, 0.67) |
| 35-39.9 | 0.78 (0.64, 0.96)[1] | 1.39 (1.27, 1.53) | 1.20 (1.13, 1.27) | 1.13 (1.03, 1.24) | 1.31 (1.06, 1.62) | 1.248 (1.240, 1.255) | 1.03 (0.93, 1.14) | 0.58 (0.37, 0.91) |
| 40+ | | 1.31 (1.11, 1.55) | 1.27 (1.17, 1.39) | 1.16 (1.02, 1.31) | 0.98 (0.74, 1.29) | 1.400 (1.391, 1.409) | 1.03 (0.91, 1.17) | 0.78 (0.48, 1.29) |
| **SPTB** | | | | | | | | |
| <18.5 | 1.25 (1.14, 1.37) | 1.39 (1.27, 1.53) | 1.36 (1.27, 1.45) | 1.33 (1.18, 1.49) | 1.92 (1.32, 2.80) | 1.405 (1.391, 1.420) | 1.62 (1.33, 1.98) | 3.59 (1.80, 7.17) |
| 18.5-24.9 | 1.00 | 1.00 | 1.00 | 1.00 | 1.00 | 1.00 | 1.00 | 1.00 |
| 25-29.9 | 0.85 (0.78, 0.92) | 1.01 (0.96, 1.07) | 0.91 (0.87, 0.95) | 0.91 (0.86, 0.98) | 0.80 (0.65, 0.98) | 0.896 (0.892, 0.901) | 0.85 (0.78, 0.94) | 0.45 (0.29, 0.69) |
| 30-34.9 | 0.85 (0.74, 0.97) | 1.09 (1.01, 1.18) | 0.90 (0.84, 0.96) | 0.85 (0.77, 0.93) | 1.06 (0.84, 1.34) | 0.891 (0.885, 0.897) | 0.72 (0.64, 0.81) | 0.38 (0.23, 0.63) |
| 35-39.9 | 0.71 (0.57, 0.89)[1] | 1.08 (0.95, 1.22) | 0.87 (0.78, 0.97) | 0.84 (0.74, 0.96) | 1.14 (0.84, 1.56) | 0.862 (0.855, 0.870) | 0.75 (0.64, 0.88) | 0.66 (0.38, 1.17) |
| 40+ | | 1.18 (0.95, 1.47) | 0.88 (0.77, 1.03) | 0.87 (0.73, 1.03) | 0.88 (0.59, 1.30) | 0.824 (0.815, 0.833) | 0.68 (0.55, 0.83) | 0.61 (0.30, 1.22) |
| **MPTB** | | | | | | | | |
| <18.5 | 0.93 (0.70, 1.25) | 1.14 (1.00, 1.30) | 1.25 (1.17, 1.34) | 1.34 (1.15, 1.56) | 1.40 (1.18, 1.66) | 1.191 (1.176, 1.206) | 1.36 (1.08, 1.70) | 1.65 (0.57, 4.81) |
| 18.5-24.9 | 1.00 | 1.00 | 1.00 | 1.00 | 1.00 | 1.00 | 1.00 | 1.00 |
| 25-29.9 | 0.80 (0.63, 1.00) | 1.30 (1.22, 1.38) | 1.14 (1.10, 1.18) | 1.18 (1.09, 1.28) | 1.10 (1.02, 1.18) | 1.176 (1.170, 1.183) | 1.03 (0.94, 1.13) | 0.69 (0.44, 1.08) |
| 30-34.9 | 1.14 (0.82, 1.57) | 1.59 (1.46, 1.72) | 1.32 (1.26, 1.39) | 1.51 (1.37, 1.66) | 1.38 (1.27, 1.51) | 1.435 (1.426, 1.434) | 1.14 (1.03, 1.27) | 0.64 (0.39, 1.04) |
| 35-39.9 | 1.28 (0.81, 2.02)[1] | 1.87 (1.64, 2.12) | 1.48 (1.37, 1.59) | 1.62 (1.43, 1.84) | 1.73 (1.55, 1.92) | 1.685 (1.672, 1.698) | 1.31 (1.15, 1.49) | 0.52 (0.26, 1.06) |
| 40+ | | 1.51 (1.19, 1.92) | 1.58 (1.42, 1.75) | 1.63 (1.37, 1.92) | 1.42 (1.24, 1.63) | 2.049 (2.032, 2.065) | 1.39 (1.18, 1.62) | 1.07 (0.56, 2.06) |

1. BMI ≥35 $kg/m^2$ as too few individuals with BMI>40 in this dataset
2. These ORs given to 3 decimal places to distinguish limits from point estimate (not possible with 2dp)

Supplementary Table S17: Adjusted odds ratios (95% CI) for any preterm, spontaneous preterm and medically indicated preterm birth by BMI category excluding multiple births

| **Any PTB** | | | | | | | | |
|---|---|---|---|---|---|---|---|---|
| BMI category | CPP, USA | Norwegian birth registry | Danish linked data | CPRD, UK | South Australian BEBOLD | US vital statistics | SAIL Databank, UK | Bradford, UK |
| <18.5 | 1.23 (1.13, 1.34) | 1.34 (1.23, 1.45) | 1.33 (1.27, 1.40) | 1.35 (1.22, 1.49) | 1.53 (1.36, 1.71) | 1.323 (1.312, 1.334) | 1.49 (1.27, 1.74) | 2.99 (1.64, 5.46) |
| 18.5-24.9 | 1.00 | 1.00 | 1.00 | 1.00 | 1.00 | 1.00 | 1.00 | 1.00 |
| 25-29.9 | 0.82 (0.76, 0.89) | 1.12 (1.07, 1.17) | 1.03 (1.00, 1.06) | 1.00 (0.95, 1.06) | 0.95 (0.90, 1.00) | 1.032 (1.028, 1.036 | 0.93 (0.87, 0.99) | 0.53 (0.38, 0.74) |
| 30-34.9 | 0.80 (0.71, 0.91) | 1.31 (1.24, 1.39) | 1.11 (1.07, 1.16) | 1.09 (1.02, 1.16) | 1.09 (1.02, 1.16) | 1.155 (1.149, 1.160) | 0.92 (0.85, 1.00) | 0.46 (0.32, 0.66) |
| 35-39.9 | 0.73 (0.60, 0.89)[1] | 1.41 (1.28, 1.55) | 1.19 (1.12, 1.27) | 1.09 (0.99, 1.20) | 1.23 (1.13, 1.34) | 1.260 (1.253, 1.268) | 1.03 (0.92, 1.14) | 0.57 (0.36, 0.91) |
| 40+ | | 1.34 (1.13, 1.59) | 1.29 (1.18, 1.41) | 1.14 (1.00, 1.29) | 1.05 (0.94, 1.17) | 1.424 (1.414, 1.433) | 1.05 (0.92, 1.19) | 0.74 (0.43, 1.28) |
| **SPTB** | | | | | | | | |
| <18.5 | 1.26 (1.16, 1.38) | 1.43 (1.30, 1.58) | 1.38 (1.28, 1.47) | 1.35 (1.20, 1.51) | 1.63 (1.41, 1.88) | 1.410 (1.396, 1.425) | 1.63 (1.33, 2.00) | 3.58 (1.80, 7.12) |
| 18.5-24.9 | 1.00 | 1.00 | 1.00 | 1.00 | 1.00 | 1.00 | 1.00 | 1.00 |
| 25-29.9 | 0.82 (0.76, 0.89) | 1.01 (0.95, 1.07) | 0.91 (0.87, 0.95) | 0.91 (0.85, 0.97) | 0.82 (0.76, 0.88) | 0.897 (0.892, 0.902) | 0.85 (0.77, 0.94) | 0.42 (0.27, 0.66) |
| 30-34.9 | 0.78 (0.68, 0.90) | 1.10 (1.02, 1.20) | 0.89 (0.83, 0.95) | 0.82 (0.74, 0.90) | 0.86 (0.78, 0.94) | 0.894 (0.888, 0.900) | 0.71 (0.63, 0.80) | 0.37 (0.22, 0.62) |
| 35-39.9 | 0.68 (0.55, 0.85)[1] | 1.08 (0.94, 1.23) | 0.87 (0.78, 0.97) | 0.81 (0.71, 0.93) | 0.84 (0.73, 0.95) | 0.867 (0.858, 0.875) | 0.74 (0.63, 0.88) | 0.62 (0.35, 1.11) |
| 40+ | | 1.19 (0.95, 1.49) | 0.91 (0.79, 1.06) | 0.84 (0.70, 1.01) | 0.74 (0.62, 0.88) | 0.833 (0.825, 0.843) | 0.69 (0.56, 0.85) | 0.62 (0.29, 1.32) |
| **MPTB** | | | | | | | | |
| <18.5 | 0.96 (0.71, 1.28) | 1.16 (1.14, 1.51) | 1.29 (1.20, 1.38) | 1.36 (1.16, 1.59) | 1.39 (1.16, 1.66) | 1.197 (1.182, 1.213) | 1.32 (1.04, 1.67) | 1.63 (0.57, 5.46) |
| 18.5-24.9 | 1.00 | 1.00 | 1.00 | 1.00 | 1.00 | 1.00 | 1.00 | 1.00 |
| 25-29.9 | 0.76 (0.60, 0.96) | 1.29 (0.92, 1.05) | 1.15 (1.11, 1.20) | 1.17 (1.08, 1.27) | 1.12 (1.04, 1.21) | 1.194 (1.188, 1.202) | 1.01 (0.92, 1.11) | 0.71 (0.45, 0.74) |
| 30-34.9 | 1.05 (0.76, 1.47) | 1.65 (0.93, 1.10) | 1.32 (1.26, 1.40) | 1.55 (1.41, 1.71) | 1.40 (1.28, 1.54) | 1.470 (1.460, 1.480) | 1.15 (1.03, 1.28) | 0.60 (0.36, 0.66) |
| 35-39.9 | 1.07 (0.67, 1.72)[1] | 1.95 (0.88, 1.15) | 1.50 (1.38, 1.62) | 1.58 (1.38, 1.81) | 1.76 (1.57, 1.97) | 1.740 (1.727, 1.754) | 1.32 (1.15, 1.51) | 0.54 (0.26, 0.91) |
| 40+ | | 1.58 (0.58, 0.95) | 1.64 (1.47, 1.83) | 1.63 (1.37, 1.95) | 1.47 (1.26, 1.70) | 2.137 (2.119, 2.155) | 1.42 (1.21, 1.68) | 0.94 (0.45, 1.28) |

1. BMI ≥35 kg/m$^2$ as too few individuals with BMI>40 in this dataset
2. These ORs given to 3 decimal places to distinguish limits from point estimate (not possible with 2dp)

Supplementary Figure S2: Meta-analysis of association of pre-pregnancy BMI with any PTB, SPTB and MPTB including and omitting CPP

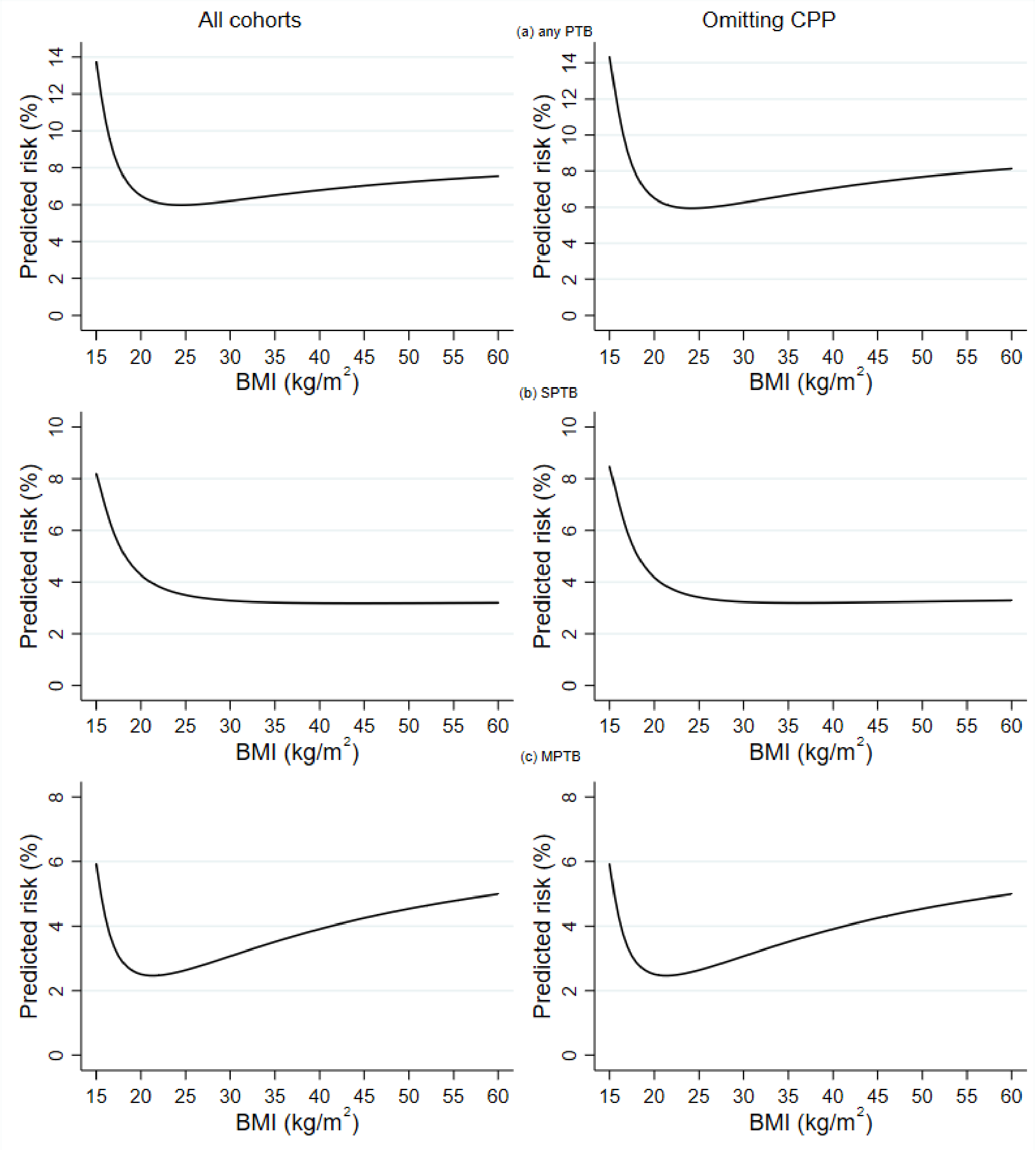
